# Epigenetic mediation may explain intergenerational associations between maternal lifestyle and children’s birth weight - Findings from the NorthPop prospective birth cohort

**DOI:** 10.1101/2025.06.01.25328723

**Authors:** Kushan De Silva, Richard Lundberg Ulfsdotter, Stina Bodén, Marie-Therese Vinnars, Patrik Ryden, Christina E. West, Magnus Domellöf, Sophia Harlid

## Abstract

**Background:** Epigenetic alterations during fetal development have been proposed as key factors explaining associations between maternal lifestyle during pregnancy and later health outcomes in the offspring, pertaining to the developmental origin of health and disease (DOHAD) hypothesis.

**Objectives:** To assess the association of maternal lifestyle with offsprings’ birth weight and underlying epigenetic mediatory mechanisms in the NorthPop prospective birth cohort.

**Methods:** A three-step analytic pipeline was applied. In 722 mother-child pairs, overall associations between 10 maternal lifestyle factors and the offspring’s standardized birth weight were first evaluated by multiple linear regression. Three high dimensional mediation methods (HDMA, HIMA, and HIMA2) were then applied on the beta methylation matrix to identify candidate CpG mediators in cord blood driving the significant overall associations. Finally, robust- and ordinary least squares-regression-based classical mediation, including single- and multiple-(parallel and serial) mediator models were assessed.

**Results:** Gestational weight gain (GWG) (β-adj = 0.03; p = 2×10^-5^) and maternal BMI at the beginning of pregnancy (β-adj = 0.036; p = 1×10^-4^) were significantly associated with the offspring’s standardized birth weight. High dimensional mediation analyses identified pooled sets of four (cg19242268; cg08461903; cg14798382; cg21516291) and five (cg17040807; cg19242268; cg26552621; cg04457572; cg06457011) candidate CpG mediators related to GWG and BMI at the beginning of pregnancy, respectively. For both exposures, classical mediation analyses revealed a range of significant single- and multiple (both serial and parallel) mediator models via both robust- and OLS-regression based approaches. These indicated the likely presence of individual-, causally linked multiple-, and causally independent multiple mediatory pathways underlying the two significant overall associations.

**Conclusions:** Our findings support the hypothesis that neonatal health effects related to maternal lifestyle may be partly mediated by epigenetic alterations. Findings also suggest the possible involvement of multiple DNA methylation sites via various mediatory pathways.

## INTRODUCTION

Epidemiological studies that lend support for the impact of maternal lifestyle on offspring’s life course health are mounting [1–3]. Historical cohorts have indicated intergenerational transmission of adverse health outcomes from pregnant women exposed to extreme conditions such as famines [4, 5]. Current epidemiological evidence suggests that a broad spectrum of maternal lifestyle factors including diet, sedentary behavior, smoking, alcohol consumption, and obesity may have long lasting effects on the offspring’s health [1–3, 6, 7]. For example, maternal smoking during pregnancy may influence fetal development through mechanisms including in utero hypoxia, nicotine-induced uteroplacental blood flow diminution, and placental toxicity [8] while maternal obesity could increase the risk of childhood obesity and overall cardiometabolic health [1–5]. Maternal diet during pregnancy likely affects nutrient availability to the fetus, whereas regular physical activity during pregnancy may optimize maternal health through mechanisms such as blood glucose homeostasis, healthy weight management, and enhanced cardiovascular fitness, leading to improved fetoplacental circulation and reduced risk of preterm birth [6, 7].

Birth weight is a multifaceted indicator of neonatal health reflecting the prenatal environment, nutritional status, fetal growth, and potential risks for both immediate and long-term health outcomes [9, 10]. Low birth weight is known to increase perinatal morbidity and mortality and is associated to poor cardiometabolic health in adulthood [11]. On the other hand, higher birth weights have been linked to elevated risks of obesity and type 2 diabetes later in life [12]. Notably, maternal behaviors such as diet, smoking, stress, and physical activity have also been associated with offspring’s birth weight [9].

Although mechanisms are still poorly understood, the link between maternal lifestyle and offsprings’ health outcomes is thought to be, at least partly, driven by developmental programming mediated through epigenetic modifications [13]. The concept of developmental programming is underpinned by heightened sensitivity of the developing fetus and the intrauterine environment to external stressors. Maternal metabolic disruptions may induce sustained genetic, phenotypic, and physiologic adaptations in the developing fetus, leading to lasting effects on its future health postnatally [14]. Epigenetic modifications, which entail the modulation of gene expression without altering the original DNA sequence, encompass multiple mechanisms including, histone acetylation, RNA modifications and DNA methylation. In epidemiological studies, DNA methylation is the mechanism that has been most thoroughly studied as it can be readily assessed at a large scale and previous work has supported the theory of epigenetics providing a modifiable link between maternal lifestyle and childhood health risks. One example includes a lifestyle intervention in pregnant women with obesity which was found to impact cord blood DNA methylation, which also associated to body composition in the offspring [15]. The primary aim of the current study was therefore to assess associations between maternal lifestyle and offsprings’ birth weight and evaluate underlying epigenetic mediatory mechanisms.

## MATERIALS AND METHODS

### Study population

The NorthPop Birth Cohort Study (NorthPop) is an ongoing population-based, prospective birth cohort conducted in Västerbotten county, Northern Sweden. It includes an extensive longitudinal database and a biobank. NorthPop aims to include 10,000 pregnant women and follow their children through birth until 7 years of age (https://www.umu.se/en/research/infrastructure/northpop/). With prospectively collected, lifestyle-related information of pregnant women, epigenetic measures in cord blood, and follow-up information of children at birth being available, the NorthPop cohort provides a unique opportunity to assess not only the association between maternal lifestyle and the offspring’s health but also associated putative epigenetic mediatory mechanisms.

### Study sample

A sample of 722 mother–child pairs from the NorthPop cohort, with cord blood DNA methylation measured at birth, were included in this study. Participating mothers were selected based on previous parity (primipara single-birth mothers or multiparous twin or triplet mothers) and sample availability. Eligible pregnant women were recruited during the years 2016 – 2020, from the University Hospital of Umeå catchment area at the time of their routine ultrasound examination at gestational week 14-24. Informed consent was given by all participating women and their partner. Web-based questionnaires were administered to the participating women at multiple times during and after the pregnancy. The first questionnaire was administered during gestational week 14-24, to collect information on socioeconomic status and medical history. Details on lifestyle during pregnancy including diet, physical activity, and stress, were collected through questionnaires provided at gestational week 26-34. Questions about the woman’s health during pregnancy and the health of the newborn were included in a questionnaire sent four months postpartum.

### Exposures and outcome

Ten maternal lifestyle-related exposures were originally included in the study, with details provided in **Supplementary material 1**. These comprised physical activity, stress, six different diet-related exposures, gestational weight gain (GWG) and body mass index (BMI) at the beginning of pregnancy. The outcome, birth weight, was obtained from The Swedish Pregnancy Register [16] and standardized using the latest published intrauterine growth reference ranges for estimated fetal weight applicable to Sweden [17].

### DNA methylation data

Cord blood buffy coat DNA samples from the children were bisulphite treated and analyzed for methylation using the Infinium MethylationEPIC BeadChip (Illumina). DNA quality control, pre-processing, processing, and output data quality control were performed at the SNP&SEQ Technology Platform, Uppsala, Sweden, part of the National Genomics Infrastructure (NGI) Sweden and Science for Life Laboratory.

The methodological workflow consisted of three steps as outlined below and presented in

### Supplementary material 2

#### Step 1

We applied multiple linear regression to assess overall associations between maternal lifestyle-related exposures (n = 10) and offsprings’ standardized birth weight. Directed acyclic graphs (DAGs) were drawn a priori to determine covariates to be included in the analysis of each exposure-outcome association, using the “dagitty” R package [18]. Based on DAGs, the multiple regression modelling the association between maternal BMI at the beginning of pregnancy and standardized birth weight was adjusted for maternal age, maternal education, maternal country of birth, and maternal smoking during pregnancy. All other overall association analyses were adjusted for maternal age, maternal BMI at the beginning of pregnancy, maternal education, maternal country of birth, and maternal smoking during pregnancy. Missing data were excluded from the multiple regression analyses. The maternal exposures significantly associated with offsprings’ standardized birth weight as per Step 1, were the focus in subsequent downstream analyses.

#### Step 2

An account of the methylation data processing and the analytic pipeline is provided in **Supplementary material 3**. A DNA methylation expression matrix with beta values produced by the processing pipeline detailed in **Supplementary material 3** was used for high dimensional mediation analyses in Step 2. We applied three high dimensional mediation methods amenable for DNA methylation data to identify candidate CpG mediators that drive the significant overall associations observed in Step 1. These methods represent recent developments in epigenetic mediation analysis which strive to overcome high dimensionality by a two-step procedure. Briefly, the initial sure independence screening (SIS) step is followed by a subsequent variable selection step such as de-sparsified LASSO to further reduce dimensions. The ultimate statistical testing is performed on a low dimensional feature space which has both survived SIS (in step 1) and been identified by the de-biased LASSO (in step 2) to determine significant mediators [19]. The three methods have been described in **Supplementary material 4**. All three methods entail penalized regression to estimate mediator-specific contributions [19]. Of these, the “HDMA” [20] and “HIMA” [21] methods were deployed using the “HDMED” R package [19] while the “HIMA2” [22] method was deployed through the “HIMA” R package [21].

Covariates determined by DAGs and cord blood cell type proportion estimates were included in all high dimensional mediation analyses. A false discovery rate (FDR) adjusted p-value threshold < 0.05 was applied to further filter the set of CpG sites selected by each high dimensional mediation method and determine candidate CpG mediators. Findings from each method were merged to produce the pooled set of candidate CpG mediators. Classical mediation analyses were performed on these candidate CpG mediators in Step 3.

#### Step 3

We assessed both robust-regression based bootstrap mediation models and OLS-regression-based bootstrap mediation models via the ‘robmed’ R package [23]. DNA methylation expression beta values of the candidate CpG mediators produced by the processing pipeline detailed in **Supplementary material 3** was used for classical mediation analyses in Step 3. With respect to each significant overall association identified in Step 1, we analyzed single mediator models, multiple serial mediator models (assuming causal dependence between multiple mediators), and parallel mediator models (assuming causal independence between multiple mediators). All analyses were adjusted for DAG-based covariates. As the number of mediatory pathways combinatorially increase in serial models quickly growing in complexity, the ‘robmed’ package allows only a maximum of three mediators in serial multiple mediation analyses. We determined the three CpG mediators to be included in serial mediation analyses, based on the results from single mediation assessments.

Finally, we searched the CpG mediators identified by the present study on the MRC-IEU catalog of epigenome-wide association studies (EWAS Catalog) [24] to uncover any consistent findings reported in previous studies.

## RESULTS

General characteristics of the maternal – offspring paired cohorts analyzed in the present study are summarized in **Table 1**.

**Table 1.** General characteristics of maternal- and offspring samples in the NorthPop prospective birth cohort analyzed in the present study.

| Maternal cohort |  | Offspring cohort |  |
| --- | --- | --- | --- |
| Characteristic<sup>a</sup> | N = 702 | Characteristic<sup>a</sup> | N = 722 |
| Maternal age (years)<sup>b</sup> | 29.9 (4.1) | Birth weight (grams) | 3449 (526.2) |
| Previous parity |  | Standardized birth weight (z-score) | -0.42 (0.04) |
| 0 | 682 | Birth length (cm) | 50.0 (2.3) |
| ≥ 1 | 20 | Gestational age at birth (weeks) | 39.8 (1.7) |
| Physical activity<sup>c,d</sup> | 9.4 (4.1) | Apgar |  |
| Less, n | 434 | 1-minute score | 8.4 (1.4) |
| More, n | 227 | 5-minute score | 9.0 (0.9) |
| Missing | 41 | 10-minute score | 9.4 (0.7) |
| Stress<sup>e,f</sup> | 1.9 (2.4) | Missing |  |
| No, n | 515 | Sex, n |  |
| Yes, n | 168 | Female | 334 |
| Missing, n | 19 | Male | 388 |
| DDS<sup>g</sup> | 20.7 (5.3) | Delivery Mode, n |  |
| DDS-eadj<sup>h</sup> | 21.8 (4.7) | Vaginal | 610 |
| DII<sup>i</sup> | -1.2 (1.9) | Cesarean | 112 |
| MDS<sup>j</sup> | 4.5 (1.7) | Year of birth, n |  |
| HNFI<sup>k</sup> | 2.5 (1.4) | 2016 | 41 |
| Total energy intake (kcal/d) | 2275 (777) | 2017 | 183 |
| Dietary CO<sub>2</sub> e/DCP | 1560.9 (1023.7) | 2018 | 240 |
| GWG<sup>l</sup>(kg) | 15.2 (5.6) | 2019 | 180 |
| BMI<sup>m</sup> (kg/m<sup>2</sup>) | 24.5 (4.1) | 2020 | 78 |
| <18, n | 5 |  |  |
| 18 - 24.9, n | 439 |  |  |
| ≥ 25, n | 238 |  |  |
| Missing | 20 |  |  |
| Smoking<sup>n</sup>, n |  |  |  |
| No | 681 |  |  |
| Yes | 2 |  |  |
| Missing | 19 |  |  |
| Country of birth<sup>o</sup>, n |  |  |  |
| Sweden | 630 |  |  |
| Other | 62 |  |  |
| Missing | 10 |  |  |
| Educational level<sup>o</sup>, n |  |  |  |
| 9 year primary school | 11 |  |  |
| Upper secondary education | 164 |  |  |
| University or university college | 516 |  |  |
| Missing | 11 |  |  |

a – reported as mean (SD) unless otherwise specified; b – mother's age at delivery; c – validated questionnaire-based index; d - score < 11 is 'less', score ≥ 11 is 'more'; e - based on the General Health Questionnaire – 12 items (GHQ-12); f - score ≥ 3 is 'Yes', score < 3 is 'No'; g - based on 40 food items; h - based on 40 food items and adjusted for total energy intake; i - based on 30 of the 45 food items used in the original study; j - an adapted version based on 8 food items; k - based on 6 food groups typically consumed in Nordic countries; l - self-reported at 4 months postpartum, in kg; m - calculated at the beginning of pregnancy; n - self-reported at gestational age 26 weeks; o - self-reported at gestational age 14-24 weeks
BMI – body mass index; DDS – diet diversity score; Dietary CO<sub>2</sub> e/DCP – dietary CO<sub>2</sub> emission/dietary carbon footprint; DII – diet inflammatory index; GDM – gestational diabetes mellitus; GWG – gestational weight gain; HNFI – healthy Nordi food index; IQR – inter-quartile range; MDS – mediterranean diet score; NA – number of missing data; SD – standard deviation

Results from DAGs analysis that determined the covariates to be included in overall association analyses are presented in **Supplementary material 5**. In the adjusted linear regression models, two maternal lifestyle markers, gestational weight gain (GWG) and BMI at the beginning of pregnancy, were significantly associated with offspring’s standardized birth weight (β_GWG_ = 0.03; 95% CI = 0.02 - 0.04 and β_BMI_ = 0.036; 95% CI = 0.019 - 0.054) (**Table 2**).

**Table 2:** Overall associations between maternal lifestyle markers and offspring’s standardized birth weight in the NorthPop prospective birth cohort as per simple- and multiple-linear regression.

| Maternal lifestyle factor | β-unadj | 95% CI of β-unadj | *p*-value | β-adj | 95% CI of β-adj | *p*-value |
| --- | --- | --- | --- | --- | --- | --- |
| GWG<sup>a</sup> | 0.034 | 0.021 to 0.047 | 6e-07 | 0.031 | 0.021 to 0.042 | 2e – 05 |
| Physical activity (continuous)<sup>a</sup> | -0.0011 | -0.0187 to 0.0167 | 0.91 | -0.0009 | -0.0192 to 0.0178 | 0.92 |
| More physical activity (Ref. = Less)<sup>a</sup> | -0.026 | -0.179 to 0.127 | 0.74 | 0.021 | -0.134 to 0.176 | 0.79 |
| BMI at the beginning of pregnancy<sup>b</sup> | 0.039 | 0.02 to 0.055 | 3e-05 | 0.036 | 0.019 to 0.054 | 1e – 04 |
| Total energy intake<sup>a</sup> | 0.00003 | -0.00007 to 0.00011 | 0.58 | 0.00005 | -0.00004 to 0.00014 | 0.28 |
| Stress (continuous)<sup>a</sup> | 0.017 | -0.013 to 0.047 | 0.27 | 0.007 | -0.024 to 0.039 | 0.65 |
| More stress (Ref = Less)<sup>a</sup> | 0.027 | -0.138 to 0.193 | 0.75 | -0.026 | -0.196 to 0.143 | 0.76 |
| Diet diversity score<sup>a</sup> | 0.003 | -0.011 to 0.017 | 0.69 | 0.009 | -0.005 to 0.023 | 0.22 |
| Dietary CO<sub>2</sub> emission/carbon footprint<sup>a</sup> | 0.00001 | -0.00006 to 0.00008 | 0.69 | 0.000008 | -0.00006 to 0.00008 | 0.83 |
| Diet inflammatory index<sup>a</sup> | 0.025 | -0.013 to 0.063 | 0.20 | 0.007 | -0.032 to 0.046 | 0.73 |
| Mediterranean diet score<sup>a</sup> | -0.038 | -0.081 to 0.005 | 0.09 | -0.017 | -0.062 to 0.027 | 0.45 |
| Healthy Nordic food index<sup>a</sup> | 0.022 | -0.03 to 0.075 | 0.40 | 0.042 | -0.011 to 0.096 | 0.12 |

a) adjusted for maternal age, maternal BMI at the beginning of pregnancy, maternal education, maternal country of birth, maternal smoking during pregnancy b) adjusted for maternal age, maternal education, maternal country of birth, maternal smoking during pregnancy
BMI – body mass index; CI – confidence interval; GWG – gestational weight gain; β-adj – effect size measured as the multiple linear regression coefficient; $\beta$ -unadj – effect size measured as the simple linear regression coefficient

The HDMA method identified 21 CpG sites mediating the association between GWG and offspring’s standardized birth weight, four of which (cg19242268; cg08461903; cg14798382; cg21516291) passed an FDR adjusted threshold of 0.05 and were selected as candidate CpG sites for classical causal mediation analysis. The HIMA method derived a set of 24 CpG sites mediating the association between GWG and offspring’s standardized birth weight, 3 of which (cg19242268; cg08461903; cg21516291) passed the FDR-adjusted threshold of 0.05 and were selected as candidate CpG sites for classical causal mediation analysis. Finally, the HIMA2 method also identified 24 CpG sites as mediating the association between GWG and the offsprings’ standardized birth weight, 2 of which (cg19242268; cg08461903) passed the FDR-adjusted threshold of 0.05 and were selected for classical causal mediation analysis. The pooled set of candidate CpG sites (n = 4) eligible for classical mediation analysis of the association between GWG and the offspring’s standardized birth weight included the same four CpG sites as captured by the HDMA method (cg19242268; cg08461903; cg14798382 and cg21516291) (**Table 3; Supplementary material 6**).

**Table 3:** Summary of results from high-dimensional mediation analysis including the candidate CpG sites selected as mediators.

| Association between GWG and offspring's z-birth weight |  |  |  |  |
| --- | --- | --- | --- | --- |
| High dimensional mediation analytic approach | CpG sites selected by high dimensional mediation approach after 2-step dimension reduction | | Candidate CpG sites selected for classical causal mediation analysis (FDR-adjusted $p < 0.05$) | |
|  | Number | Composition | Number | Composition |
| HDMA | (n = 21) | cg04968127; cg14798382; cg21516291; cg27053299; cg10660916; cg14556683; cg10178960; cg16752400; cg09247736; cg18137450; cg07002832; cg02832224; cg05779272; cg19242268; cg16402875; cg15672022; cg13131501; cg04457572; cg01940139; cg08461903; cg00154986 | (n = 4) | cg19242268; cg08461903; cg14798382; cg21516291 |
| HIMA | (n = 24) | cg04968127; cg05349624; cg21516291; cg27053299; cg10660916; cg14556683; cg10178960; cg16752400; cg05304729; cg04751761; cg09247736; cg18137450; cg02832224; cg05779272; cg19242268; cg05560494; cg16402875; cg13131501; cg12804755; cg04457572; cg22247250; cg08461903; cg00154986; cg12145085 | (n = 3) | cg19242268; cg08461903; cg21516291 |
| HIMA2 | (n = 24) | cg04968127; cg05349624; cg21516291; cg27053299; cg10660916; cg14556683; cg10178960; cg16752400; cg05304729; cg04751761; cg09247736; cg18137450; cg02832224; cg05779272; cg19242268; cg05560494; cg16402875; cg13131501; cg12804755; cg04457572; cg22247250; cg08461903; cg00154986; cg12145085 | (n = 2) | cg19242268; cg08461903 |
| Association between maternal BMI and offspring's z-birth weight |  |  |  |  |
| High dimensional mediation analytic approach | CpG sites selected by high dimensional mediation approach after 2-step dimension reduction | | Candidate CpG sites selected for classical causal mediation analysis (FDR-adjusted $p < 0.05$) | |
|  | Number | Composition | Number | Composition |
| HDMA | (n = 25) | cg04968127; cg21516291; cg23260105; cg00376553; cg17040807; cg21649604; cg16752400; cg25494075; cg08289567; cg18137450; cg02832224; cg05779272; cg19242268; cg26552621; cg18034719; cg09171931; cg13131501; cg03688987; cg04457572; cg06457011; cg14787880; cg01940139; cg08461903; cg00154986; cg12145085 | (n = 5) | cg17040807; cg19242268; cg26552621; cg04457572; cg06457011 |
| HIMA | (n = 24) | cg04968127; cg14798382; cg21516291; cg23260105; cg00376553; cg17040807; cg16752400; cg05304729; cg15482893; cg18137450; cg02832224; cg05779272; cg19242268; cg26552621; cg18034719; cg13131501; cg03688987; cg04457572; cg06457011; cg14787880; cg01940139; cg08461903; cg00154986; cg05632420 | (n = 5) | cg17040807; cg19242268; cg26552621; cg04457572; cg06457011 |
| HIMA2 | (n = 24) | cg04968127; cg14798382; cg21516291; cg23260105; cg00376553; cg17040807; cg16752400; cg05304729; cg15482893; cg18137450; cg02832224; cg05779272; cg19242268; cg26552621; cg18034719; cg13131501; cg03688987; cg04457572; cg06457011; cg14787880; cg01940139; cg08461903; cg00154986; cg05632420 | - | - |

BMI – body mass index; GWG – gestational weight gain

The same methods were used to identify CpG sites mediating the association between maternal BMI at the beginning of pregnancy and the offspring’s standardized birth weight. The HIMA, HDMA, and HIMA2 methods yielded 24, 25, and 24 CpG mediatory sites, respectively. Of these, both HDMA and HIMA output comprised the same subset of 5 candidate CpG sites that passed the FDR-adjusted threshold of 0.05 and were selected for classical causal mediation analysis (cg17040807; cg19242268; cg26552621; cg04457572; cg06457011) (**Table 3; Supplementary material 6**).

Two robust regression-based single mediator models (cg19242268; cg14798382) (**Table 4; Figure 1**) and three OLS regression-based single mediator models (cg19242268; cg14798382; cg08461903) (**Supplementary material 7; Supplementary material 8**) examining the association between GWG and offspring’s standardized birth weight were significant. All three CpG sites were also identified by the high-dimensional mediation analyses described above and included in multiple mediator models.

**Figure 1:**
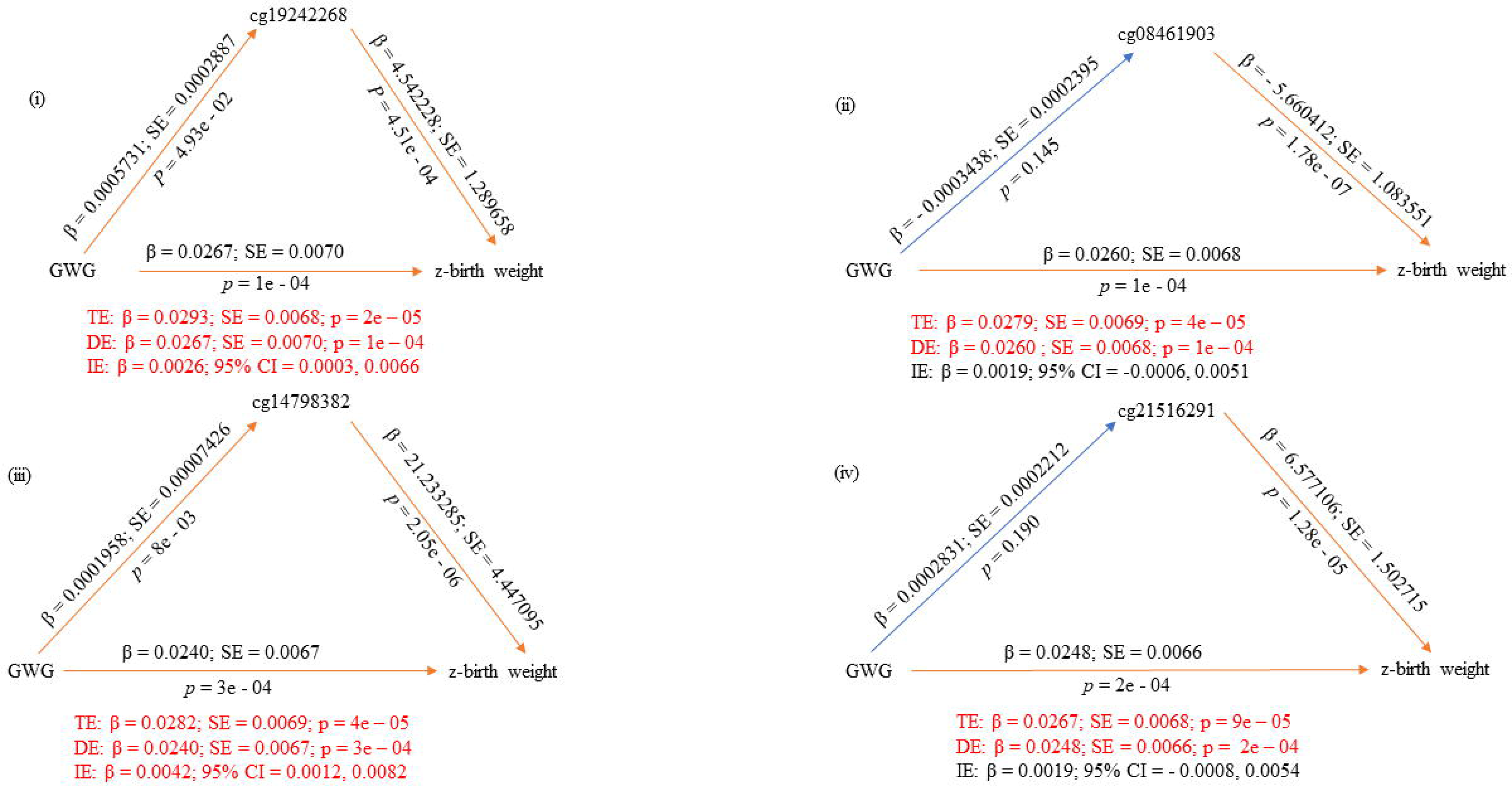
Single mediator models as per the robust bootstrapped approach with candidate CpG sites as mediators of the association between GWG and offspring’s z-birth weight.

**Table 4:**
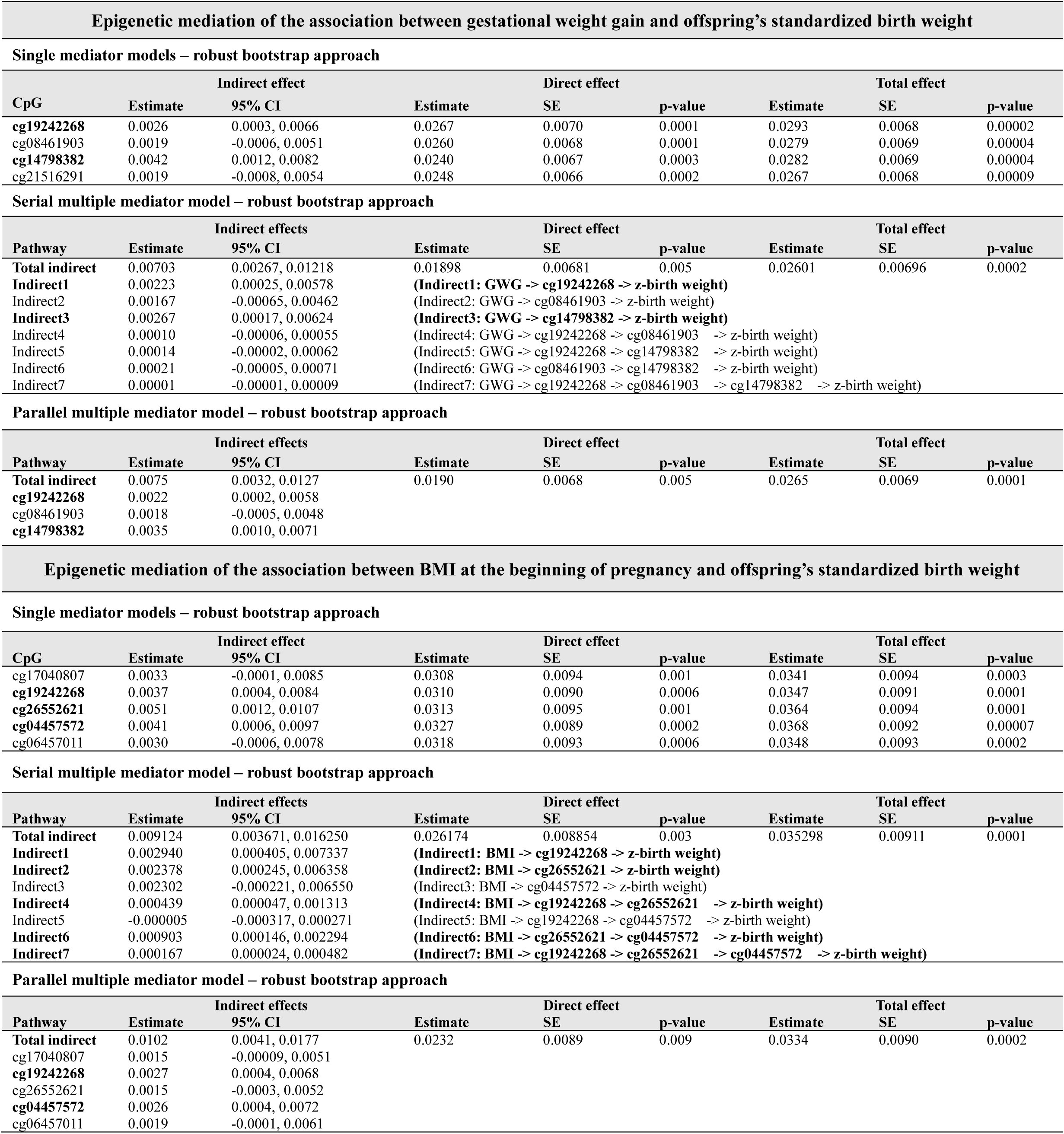
Summary of results from robust mediation analysis with candidate CpG sites selected by high dimensional mediation analysis.

When examining serial multiple mediation, we identified several significant indirect pathways, two in robust models (GWG -> cg19242268 -> z-birth weight and GWG -> cg14798382 -> z-birth weight) (**Table 4; Figure 2**) and five in OLS models (GWG -> cg19242268 -> z-birth weight, GWG -> cg08461903 -> z-birth weight, GWG -> cg14798382 -> z-birth weight, GWG -> cg19242268 -> cg14798382 -> z-birth weight and GWG -> cg08461903 -> cg14798382 -> z-birth weight) (**Supplementary material 7; Supplementary material 8**).

**Figure 2:**
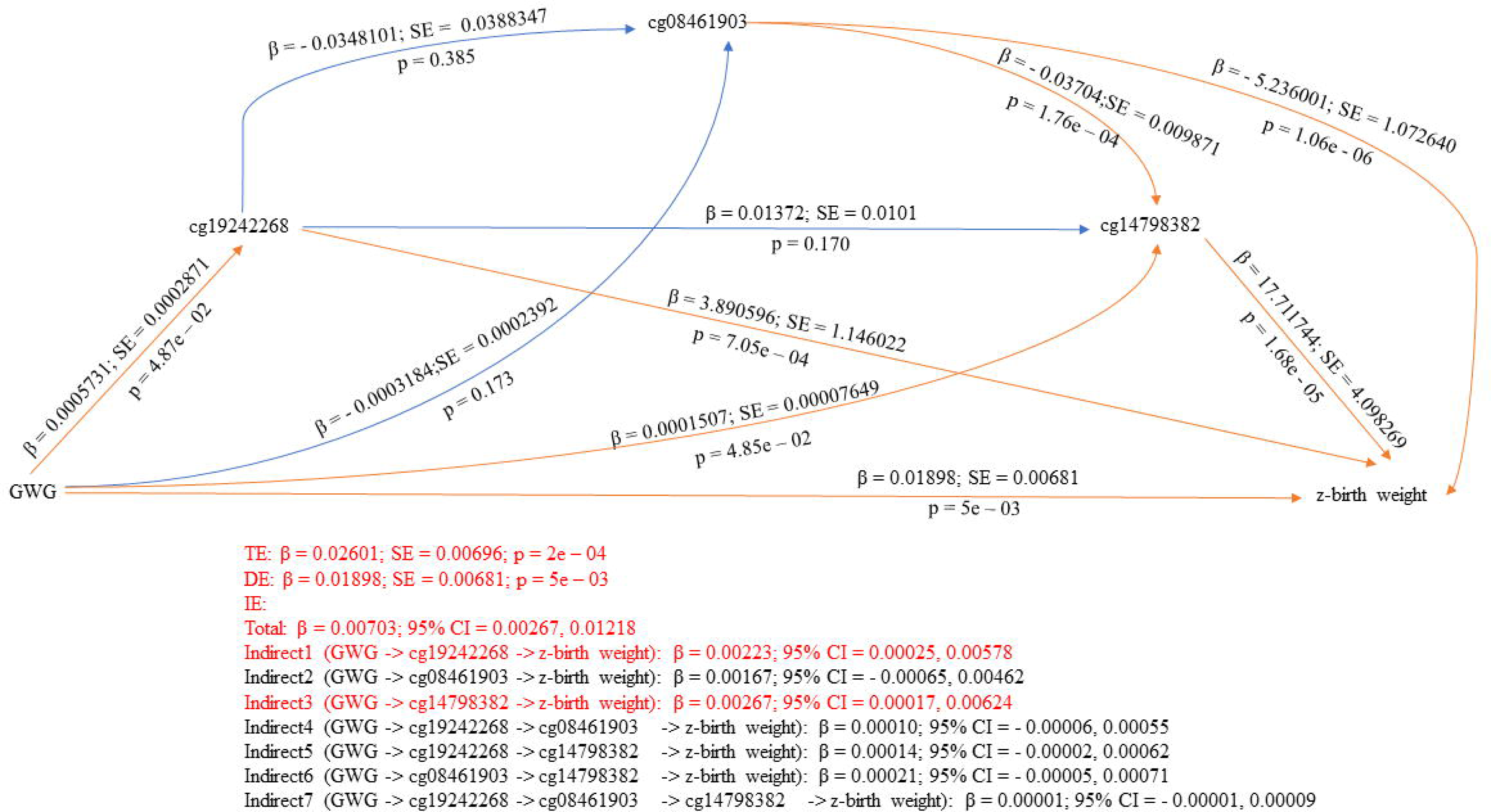
Serial multiple mediator model as per the robust bootstrapped approach with candidate CpG sites as mediators of the association between GWG and offspring’s z-birth weight.

Robust parallel multiple mediation of GWG’s association with offspring’s standardized birth weight revealed two significant indirect pathways (cg19242268 and cg14798382) (**Table 4; Figure 3**), whereas OLS parallel multiple mediation of the same association found three indirect pathways (cg19242268; cg08461903; cg14798382) (**Supplementary material 7; Supplementary material 8**).

**Figure 3:**
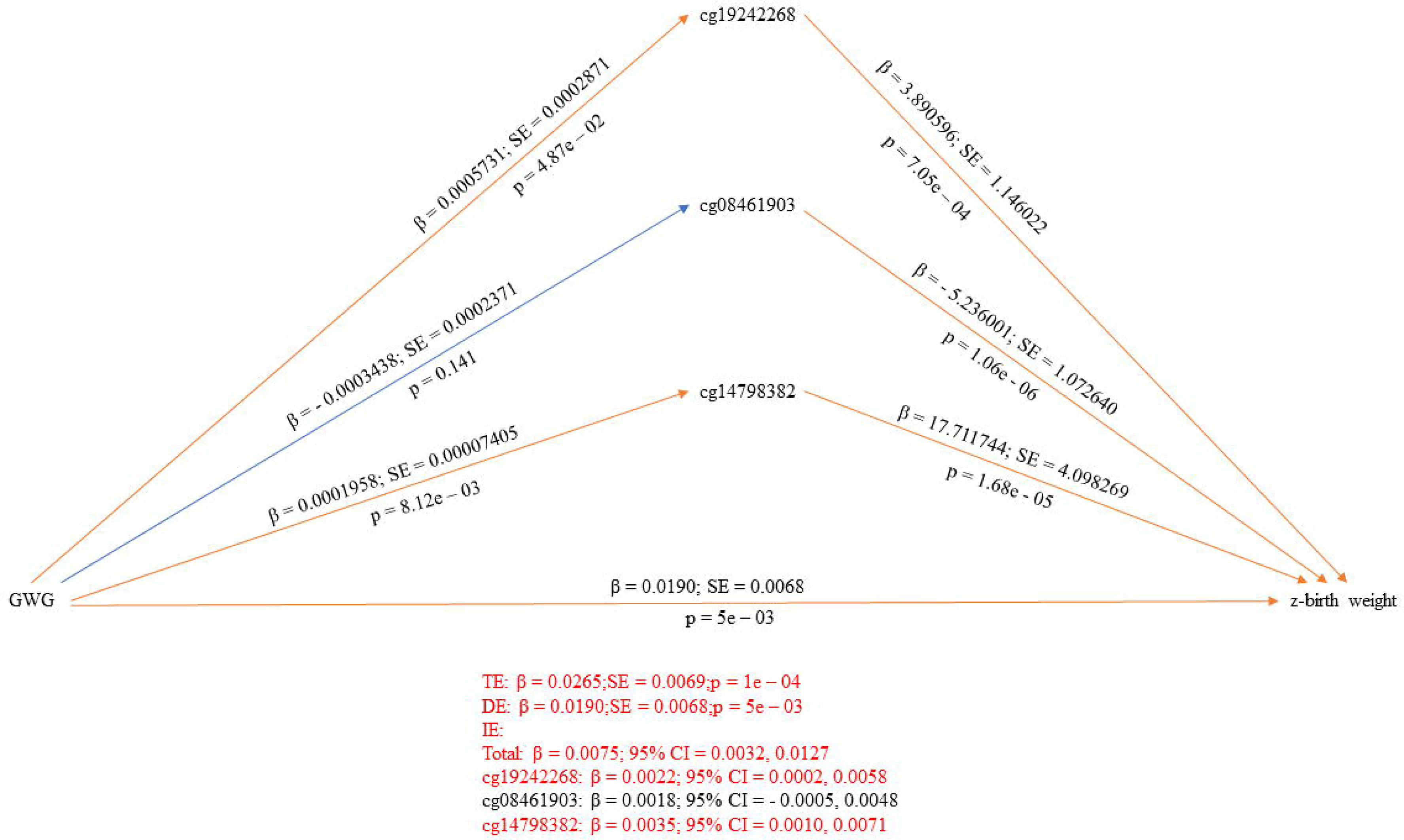
Parallel multiple mediator model as per the robust bootstrapped approach with candidate CpG sites as mediators of the association between GWG and offspring’s z-birth weight.

Three robust single mediator models (cg19242268: cg26552621; cg04457572) (**Table 4**) and all five OLS single mediator models (cg17040807; cg19242268; cg26552621; cg04457572; cg06457011) examining the association between maternal BMI and offspring’s standardized birth weight were significant (**Supplementary material 7; Supplementary material 8**). The three significant CpG sites in robust single mediator models were included in serial multiple mediation.

We observed several significant indirect pathways as per serial multiple mediation, five robust pathways (BMI -> cg19242268 -> z-birth weight; BMI -> cg26552621 -> z-birth weight; BMI -> cg19242268 -> cg26552621 -> z-birth weight; BMI -> cg26552621 -> cg04457572 -> z-birth weight; BMI -> cg19242268 -> cg26552621 -> cg04457572 -> z-birth weight) (**Table 4**) and six OLS pathways (BMI -> cg19242268 -> z-birth weight; BMI -> cg26552621 -> z-birth weight; BMI -> cg04457572 -> z-birth weight; BMI -> cg19242268 -> cg2655262 -> z-birth weight; BMI -> cg26552621 -> cg04457572 -> z-birth weight; BMI -> cg19242268 -> cg26552621 -> cg04457572 -> z-birth weight) (**Supplementary material 7; Supplementary material 8**).

In contrast, parallel robust multiple mediation revealed two significant indirect pathways (cg19242268; cg04457572) (**Table 4**) while OLS multiple parallel mediation found three significant pathways (cg19242268; cg04457572; cg06457011) (**Supplementary material 7; Supplementary material 8**) for the association between maternal BMI and offspring’s standardized birth.

In total, eight CpG sites were selected as potential mediators of associations between GWG or pregnancy BMI and birth weight (**Table 5)**. Previous studies on the association of candidate CpG sites with markers of obesity found on the EWAS Catalog are summarized in **Supplementary material 9**. A single candidate CpG site, namely, cg19242268 was found to mediate both significant overall associations, emerging significant in all single- and multiple-mediator models.

**Table 5:** Annotated details of the CpG sites selected as mediators of the association between gestational weight gain/maternal BMI at the beginning of pregnancy and offspring’s standardized birth weight.

| CpG site | Exposure(s) | CHR | Position | Gene | Gene region | Relation to island |
| --- | --- | --- | --- | --- | --- | --- |
| cg14798382 | GWG | chr19 | 16629806 | CHERP; C19orf44 | 3'UTR | N_Shore |
| cg19242268 | GWG & BMI | chr20 | 62688573 | TCEA2 | 1<sup>st</sup> Exon; 5'UTR | Island |
| cg21516291 | GWG | chr20 | 44979100 | SLC35C2 | Body | OpenSea |
| cg08461903 | GWG | chr21 | 45884825 | - | - | S_Shore |
| cg17040807 | BMI | chr17 | 74533282 | CYGB | Body | Island |
| cg26552621 | BMI | chr19 | 1271019 | C19orf23; CIRBP | TSS 1500; Body | S_Shore |
| cg04457572 | BMI | chr10 | 73303234 | CDH23 | Body | OpenSea |
| cg06457011 | BMI | chr20 | 39767490 | PLCG1 | Body | S_Shore |

BMI – body mass index; GWG – gestational weight gain, CHR - chromosome

## DISCUSSION

We identified eight potential CpG mediators that could be mapped to genes with obesogenic potential. Seven of these mediated associations between either GWG or pregnancy BMI and birth weight, whereas one, cg19242268, stood out as a potential mediator in all models. Cg19242268 is positioned in a CpG island situated in the first exon of one isoform of the gene *TCEA2* (Transcription Elongation Factor A2), the protein of which is involved in transcriptional regulation and mainly expressed in the testis and brain. Interestingly, at least one previous study identified a differentially methylated region (DMR) in cord blood associated with birthweight that overlapped with the promoter of *TCEA2* and another gene (*RP13-152O15.5*) [25], lending further support for its involvement as an important epigenetic mediator of weight.

Another CpG of interest was cg14798382 which mapped to the *CHERP* (calcium homeostasis ER protein) gene, previously shown to be involved in cellular growth and proliferation through the regulation of calcium homeostasis [26]. In another study which aimed to identify genes associated with nonalcoholic fatty liver disease, *CHERP* was shown to be strongly downregulated in afflicted individuals [27], but still not much is known about its potential involvement in disease development.

Other CpGs of interest were situated in the genes *SLC35C2* (cg21516291), *CYGB* (cg17040807), *CIRBP* (cg26552621) and *PCLG1* (cg06457011). The *SCL35C2* gene regulates glycosylation – an essential post-translational modification process important for multiple biological processes, including embryonic development [28]. *CYGB* is essential for regulation of adipogenesis, inflammation, blood pressure, and oxidative stress response [29, 30], *CIRBP* is involved in regulation of glucose metabolism, adipose tissue function, and inflammation [31, 32] and finally, *PLCG1* is involved in insulin signaling, leptin signaling, and the regulation of adipose tissue functions [33]. Taken together, mediatory CpG sites found by the present study are situated close to multiple genes capable of elevating an individual’s obesogenic risk through diverse functional pathways.

Furthermore, consistent findings from existing literature on the EWAS Catalog [24] add to the biologically plausibility. A multi-ancestry meta-analysis of epigenome-wide association studies revealed that three mediators identified by the present study (cg19242268 in *TCEA2*, cg21516291 in *SLC35C2*, and cg17040807 in *CYGB*) are all previously identified DNA methylation markers of birth weight [34]. Meanwhile, a previous EWAS reported that seven of the methylation mediators revealed by the current study (cg19242268, cg21516291, cg26552621, cg04457572, cg06457011, cg14798382, cg08461903) associate with childhood growth trajectories from birth to late adolescence [35]. Moreover, two mediators in the present study (cg21516291 and cg04457572) were identified as epigenetic markers of incident type 2 diabetes by another EWAS [36]. These findings from previous EWAS studies bolster the credible link between the CpG mediators identified through the present analysis and obesity phenotypes.

Interestingly, an integrated methylome- and phenome-wide assessment of the circulating proteome revealed the associations of cg26552621 in *CIRB2* (identified as an epigenetic mediator in the present study) with obesogenic NOG protein levels and cg04457572 in *CDH23* (another epigenetic mediator in the present study) with ADIPOQ protein/adiponectin levels which regulate fat metabolism and insulin sensitivity [37]. Moreover, an integrative cross-omics analysis of DNA methylation sites of glucose and insulin homeostasis found that a third epigenetic mediator of the current study, cg06457011 in *PLCG1* was associated with fasting insulin while differential methylation explained at least 16.9% of the association between obesity and insulin [38]. Some of these obesity-associated methylation signatures have been robustly replicated across cohorts [34].

Previous large-scale observational epidemiological studies have linked maternal obesity with large-for-gestational-age offspring [39, 40] and suggested a potential causal relationship [41] and several of the methylation sites identified in our study have been connected to birth weight, obesity, and diabetes. However, information on mediatory pathways have been lacking. Using both robust and OLS regression-based multiple mediation analytic models we have been able to unravel potentially causally linked and causally independent mediatory pathways involving multiple methylation sites. We thus provide suggestive evidence that the methylation sites may exert their mediatory effects individually as well as concomitantly via complex pathways. These statistically intuitive findings together with existing literature shed light on the complex nature of epigenetic mediation that may underlie later health effects in the offspring highlighting epigenetic mediation as a likely mechanism contributing to intergenerational obesity.

The present study adopted a comprehensive analytic pipeline. It comprised the application of multiple topical high dimensional mediation methods followed by both OLS- and robust classical mediation analyses and the evaluation of single as well as multiple mediatory models. High dimensional mediation methods are especially amenable to DNA methylation data as revealed by previous birth cohort studies yielding novel findings [42–44].

According to our knowledge, this is the first and the largest prospective birth cohort study in a Northern European setting to unravel the epigenetically mediated association of two maternal lifestyle markers in pregnancy with children’s birth weight. A previous birth cohort study provided some evidence of epigenetics being involved in the intergenerational risk of obesity, however, their study population consisted of a predominantly urban, low-income ethnic minority and results might therefore be difficult to generalize for other populations [45].

Our study also has imitations. Although, our sample is of considerable size compared to most epigenetic studies reported earlier, larger cohorts may be required to gain more robust findings with higher statistical power. The presence of residual confounding may have influenced effect estimates, despite the inclusion of an array of DAG-based covariates. Causal inference cannot be drawn from observational designs and future studies are recommended to validate our findings. Furthermore, despite there being potential connections between birthweight and early childhood BMI trajectories the follow-up time in the current study was too short to study such associations. However, NorthPop aims to follow the included children until at least 7 years of age, with data being collected at the ages of 18 months, 3 years, and 7 years. With these data, weight trajectories of the offspring cohort will be analyzed in future studies. Since the temporal patterns of the association might be complex and change as the offspring grows, we will also investigate the potential epigenetic mediation of BMI at different ages of the child.

## CONCLUSIONS

We present new insights suggesting epigenetic factors as mediators of associations between maternal lifestyle and birthweight in this predominantly Northern European population. Our top findings include identification of eight CpG sites that appear to mediate associations between maternal characteristics (GWG and pregnancy BMI) and children’s birth weight. The most notable methylation site was cg19242268 in *TCEA2*, as DNA methylation of this site was involved in mediation between both characteristics and birth weight. Cord blood DNA methylation surrounding this gene has also previously been implicated as a marker of birth weight [25]. However, most importantly, we believe that our study results may increase the general understanding of intergenerational inheritance of obesity and highlights the importance of adhering to healthy lifestyle throughout the life span, with could benefit potentially transcending generations. Future studies are warranted to validate and elucidate the functional mechanisms.

## Data Availability

Data described in the manuscript, will be made available upon reasonable request pending valid ethical approval as well as approval by the NorthPop steering committee.

## Acknowledgments

We acknowledge all participating families in the NorthPop cohort; the NorthPop project team for recruitment, follow-up, and blood samplings of study participants, and the personnel at Biobanken Norr, Västerbotten county council. Methylation profiling was performed by the SNP&SEQ Technology Platform in Uppsala (www.genotyping.se). The facility is part of the National Genomics Infrastructure (NGI) Sweden and Science for Life Laboratory. The SNP&SEQ Platform is also supported by the Swedish Research Council and the Knut and Alice Wallenberg Foundation.

## SUPPLEMENTARY METERIALS

**Supplementary material 1: Details of the methods of maternal lifestyle markers development**

**Supplementary material 2: Study workflow**

**Supplementary material 3: DNA methylation data preprocessing and analysis pipeline**

**Supplementary material 4: Details of high dimensional mediation algorithms**

**Supplementary material 5: Directed acyclic graphs**

**Supplementary material 6: Details of the results of high dimensional mediation analysis**

**Supplementary material 7: Summary of results from ordinary least squares bootstrap mediation analysis with candidate CpG sites selected by high dimensional mediation analysis**

**Supplementary material 8: Single- and multiple-mediator models (graphical presentations)**

**Supplementary material 9: Findings from previous studies on the association of candidate CpG sites with markers of obesity**

## STATEMENTS & DECLARATIONS

### Author contributions

Kushan De Silva, Christina E West, Magnus Domellöf and Sophia Harlid contributed to the study conception and design. Material preparation, data collection and analysis were performed by Kushan De Silva, Richard Lundberg Ulfsdotter and Stina Bodén. The first draft of the manuscript was written by Kushan De Silva, and all authors critically reviewed and commented on the manuscript. All authors read and approved the final manuscript.

### Ethics approval

The NorthPop study was approved by the Research Ethics Committee in Umeå, Sweden, 2014/224-31.

### Consent to participate

Written informed consent was obtained from both parents.

