## supplementary materials for "Epigenetic mediation may explain intergenerational associations between maternal lifestyle and children’s birth weight - Findings from the NorthPop prospective birth cohort"

**Supplementary material 1: Details of the methods of maternal lifestyle markers development**

**Physical activity**: Physical activity during pregnancy was estimated by a questionnaire from the Swedish National Board of Health and Welfare (NBHW) based on validated categorical questions. For this purpose, the NBHW questionnaire was included as a part of the NorthPop questionnaire and sent out at gestational age 34 weeks. A total physical activity score was calculated by multiplying exercise time (scores for vigorous physical activity) by two and adding the product to the score for daily (moderate) physical activity, producing an index ranging from 3 to 19. The NBHW recommends a total score ≥ 11 to demarcate optimal physical activity and a score < 11 as inadequate.

**Stress**: The General Health Questionnaire – 12 items (GHQ-12) was administered at gestational age 26 weeks to pregnant women participating in the NorthPop study to measure their self-reported stress/psychological well-being. The GHQ-12 is a self-reported questionnaire used to examine the degree of general mental health/mental well-being in recent weeks. It comprises 12 statements half of which have been designed negative and the other half positive, in order to minimize response fix. The 12 items are rated on a 4-point scale and scored in several ways. Of these, the bimodal GHQ scoring method (0-0-1-1) and the Likert scoring method (0-1-2-3) are widely used. In this study, we used the bimodal GHQ scoring method (range: 0 – 12). As recommended, participants with a GHQ-12 score ≥ 3 were demarcated as of "reduced psychological well-being" while those with a score < 3 were assumed to be of "sufficient psychological well-being". Regardless of the estimation method, the GHQ-12 has demonstrated high validity and internal reliability in various settings including in Sweden. The instrument has also been used in previous research to investigate the psychological well-being of pregnant women.

**Diet inflammatory index**: The literature-derived dietary inflammatory index (Shivappa et al., 2014) has been designed to assess the inflammatory effect of diet in any population and independent of specific dietary assessment method. Dietary assessment in pregnant women participating in the NorthPop study was made via food frequency questionnaire (FFQ)s. The original DII has been derived from 45 food parameters associated with inflammatory markers (IL-1β, IL-4, IL-6, IL-10, TNF-α and C-reactive protein) based on contemporary scientific literature. These 45 parameters have been assigned an overall inflammatory score based on the strength of the association and the quality of the studies. For each parameter, food consumption data from eleven populations in different regions of the world have been used to calculate global mean intakes and standard deviations. An individual's exposure is calculated by subtracting the global mean intake from the reported intake of each food parameter, dividing by the global standard deviation and converting to a percentile score. The sum of these food-parameter-specific contributions yields the final DII score. The DII score could range from −8.87 (maximally anti-inflammatory to +7.98 (maximally pro-inflammatory). Intakes of 30 of the 45 food parameters assessed by the original study could be retrieved from the FFQs administered in the NorthPop study. These included total energy, total fat, saturated fat, trans fatty acids, cholesterol, monounsaturated fat, polyunsaturated fat, omega-6 fatty acids, omega-3 fatty acids, carbohydrates, protein, fiber, alcohol, caffeine, tea, folic acid, selenium, niacin, iron, zinc, thiamin, vitamin B2, vitamin B6, vitamin B12, vitamin A, vitamin E, vitamin C, vitamin D, magnesium, and β-carotene. Eugenol, garlic, ginger, onion, saffron, turmeric, flanan-3-ol, flavones, flavonols, flavonones, anthocyanidins, isoflavones, pepper, thyme/oregano and rosemary were the 15 omitted parameters. The DII of the present study was calculated as per the method described in the original study but based on the 30 aforementioned parameters (instead of 45).

**Diet diversity score**: Dietary data from the pregnant women collected through a self-reported FFQ in pregnancy was used to calculate the diet diversity (DD) score. A total of 40 food items were included to calculate a DD score based on the present dietary guidelines for pregnant women in Sweden (Norman et al., 2019). The DD score could potentially range from 0 to 40 points for an individual and was adjusted for total energy intake in kcal per day during pregnancy using the residual method. Detailed information about the development of DD score applied in this study has been published elsewhere (Bodén et al., 2023).

**Mediterranean diet score**: High intake of vegetables, legumes, fruits, nuts, seeds, cereals, and olive oil, moderately high intake of fish, low to moderate intake of dairy products, moderate intake of alcohol, and low intake of saturated fat, meat and meat products are the cornerstones of Mediterranean diet (Trichopoulou & Critselis, 2004). An adapted version of the Mediterranean diet score (MDS) previously applied to Swedish populations (Bodén et al., 2019) was used in the present study. The adapted MDS comprised three unfavorable food groups and five favorable food groups. The unfavorable food groups included dairy products, meat and meat products, and alcohol (>50 g/day). The favorable food groups included vegetables and potatoes, fruits and nuts, fish and fish products, MUFA+PUFA/SFA-ratio, and whole grain and cereals. The intake of all parameters except

MUFA+PUFA/SFA-ratio and alcohol were adjusted to daily energy intakes of 2000 kcal using the nutrient density method. Participants with missing FFQ data and energy intakes below the 1st percentile or >5000 kcal/day were excluded. For favorable parameters, a value of 1 was assigned to subjects whose consumption was higher than the study population-specific median and 0 for intakes below the median. For unfavorable parameters a value of 0 was assigned to subjects whose consumption was higher than the study population-specific median and 1 for intakes below the median, except for alcohol where participants with intakes > 50g/day were assigned 0, and 1 if ≤ 50g/day. The 8 parameter scores were summed for a MDS ranging from 0 (lowest adherence) to 8 (highest adherence).

**Healthy Nordic food index**: The healthy Nordic food index (HNFI) based on Olsen et al. (Olsen et al., 2011), considers dietary items likely to have beneficial health effects when the consumption is high and is based on six typical food groups consumed in the Nordic countries (fish, cabbages, whole grain rye, whole grain oats, apples and pears, and root vegetables). Based on the FFQ administered during pregnancy in the NorthPop cohort, a score of 0 or 1 was assigned each food group depending on whether the participant's intake was below or above the study population-specific median for that particular food group. The 6 food group scores were summed for a HNFI ranging from 0 (lowest adherence) to 6 (highest adherence).

**Total energy intake**: Dietary data collected from the pregnant women through a self-reported FFQ at gestational week 34 was linked to the Swedish food composition database administered by the Swedish National Food Agency, to obtain the nutrient contribution of each food item including energy content. By summing up the energy content of all food items reported in the FFQ, the total daily energy intake for each participant was calculated.

Dietary **CO2 emission/dietary carbon footprint**: Life cycle assessment (LCA) data from the Food Climate Database administered by Research Institutes of Sweden (RISE, 2019) were used to estimate greenhouse gas emissions (GHGE) associated with the diet of the pregnant women. GHGEs from the complete diet assessed using the NorthPop FFQ at gestational week 34 were calculated for all study participants and expressed in kg carbon dioxide equivalents (CO2e) per year.

**Gestational weight gain**: Gestational weight gain (GWG) data self-reported by NorthPop participants and collected at 4 months postpartum were used in the present study.

**BMI at the beginning of pregnancy**: Pregnant women's BMI measured at the beginning of pregnancy during their earliest visit at a maternity care centre was obtained from The Swedish Pregnancy Register .

**Supplementary material 2: Study workflow**

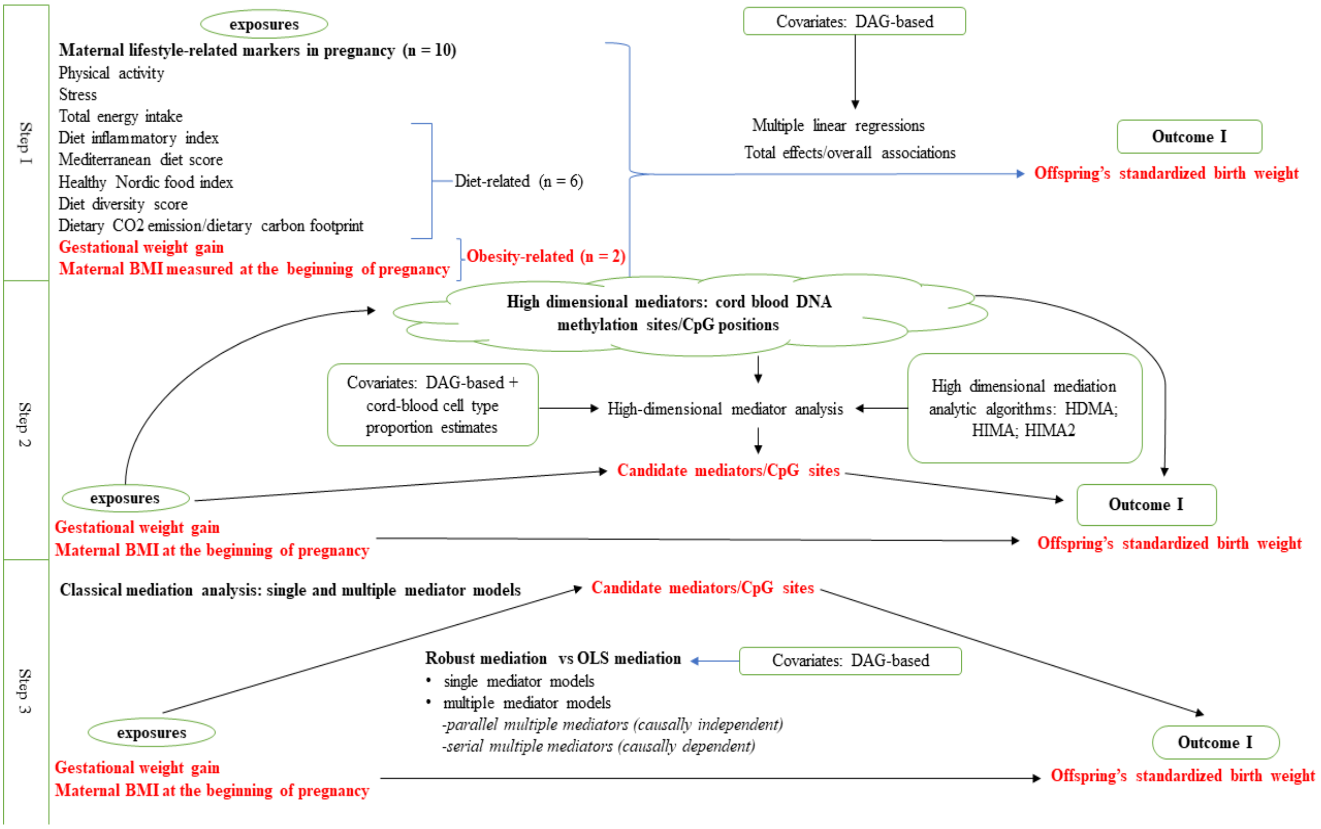

**Supplementary material 3: DNA methylation data preprocessing and analysis pipeline**

**DNA methylation data pre-processing:** Raw DNA methylation data were pre-processed using the ENmix R package. The raw DNA methylation data in IDAT format files were parsed to create an 'rgDataSet' with probe annotation. Prior to pre-processing, probes were excluded (N = 3102) if they were either SNP related, had a call-rate of P < 0.01, were outliers, or were missing in more than 20% of the samples. The threshold to identify samples with low data quality (the percentage of low-quality methylation data points across probes for each sample) was set at 5% while the threshold to identify probes with low data quality (percentage of low-quality methylation data points across samples for each probe) was specified as 20%. Non-CpG probes (n = 2650) were removed. Methylation signal intensities were background corrected using out-of-band Infinium I intensities. Quantile normalization was applied for methylation intensity values of Infinium I and Infinium II probes separately. Data were corrected for probe type bias using the regression on correlated probes (RCP) method. Methylation data were expressed as beta values at each CpG site, ranging from 0 (unmethylated) to 1 (fully methylated). As suggested by Pidsley et al. (2016), cross-reactive probes were removed. As methylation patterns can vary across cord blood cell types, we estimated cord blood cell distributions within the buffy coat fraction based on a reference dataset. Control samples (n = 8) were excluded. Known batch effects were removed using the 'ComBat' function of the 'sva' R package. We also estimated surrogate variables to account for residual confounding. This was done using intensity data for non-negative internal control probes and setting minimum percentage of variation explained by surrogate variables to 95%.

**Supplementary material 4: Details of high dimensional mediation algorithms**

**Figure 1: Conceptual framework of the high dimensional mediation model applicable to HDMA, HIMA, and HIMA2**

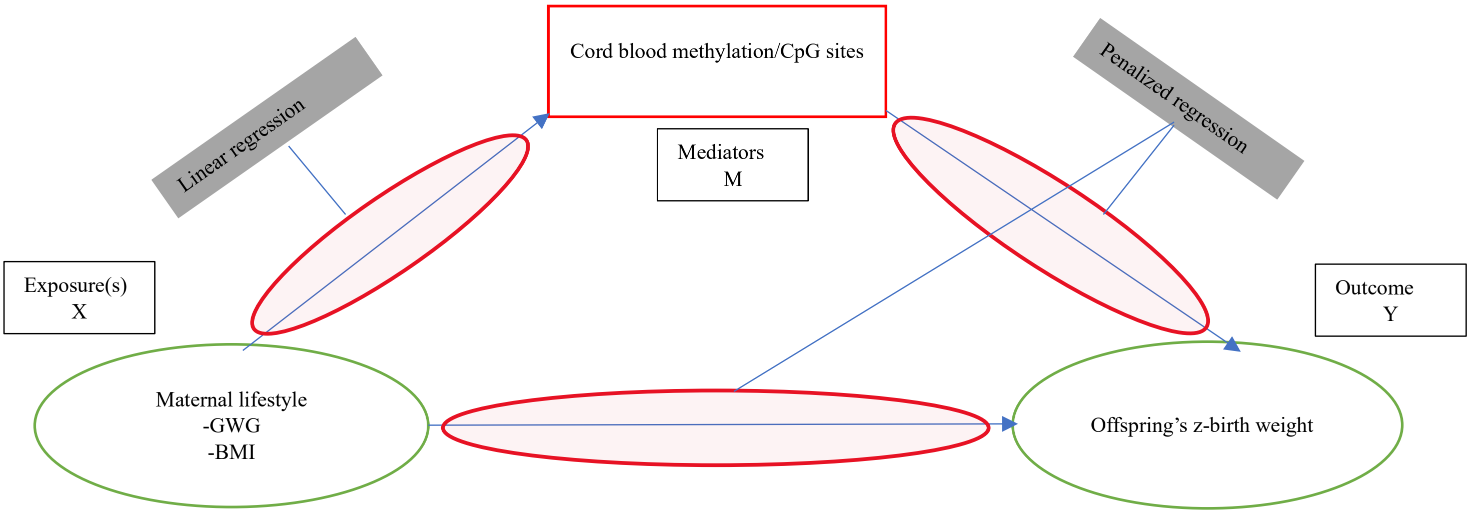


Consider the following: a sample of $N$ individuals, an exposure of interest $X_a$, a continuous outcome $Y_a$, and continuous mediators $M_a$ of the a[th] person (range of a: 1 to N). $M_a$ is a vector representing a set of R mediators $M_a^{(b)}$, whereby b ranges from 1 to R. $C_a$ is a vector of K covariates. R > 1 > N.

Outcome model:

$$E[Y_a|X_a, M_a, C_a] = \beta_a X_a + \beta_m^T M_a + \beta_c^T C_a \qquad (1)$$

Where,

- $\beta_m$ is a vector of R length in which the b[th] component, $(\beta_m)_b$, is the linear association of the b[th] mediator with $Y_a$ adjusting for the other variables
- $\beta_a$ is the association between $X_a$ and $Y_a$ adjusting for mediators and covariates

Mediator model:

$$E[M_a|X_a, C_a] = \alpha_a X_a + \alpha_c C_a \qquad (2)$$

Where,

- $\alpha_a$ is an R vector of the associations between X and each mediator, $(\alpha_a)_b$
- $\alpha_c$ is a matrix of mediator-covariate associations

The mediation contribution of an individual mediator $b$ is equivalent to $(\alpha_a)b(\beta_m)b$ (as $\sum_{b=1}^{R}(\alpha_a)b(\beta_m)b = \alpha_a^T \beta_m$)

With respect to the equations 1 and 2, assuming no residual confounding of the X-M association, no residual confounding of the X-Y association, no residual confounding of the M-Y association, that confounders of the M-Y association are not influenced by X, and no X-M interactions influencing Y:

- $\beta_a$ is the direct effect estimate of $X_a$ on $Y_a$ — (i)
- $\alpha_a^T \beta_m$ is the total indirect effect/mediatory effect of $X_a$ on $Y_a$ via $M_a$ — (ii)
- And the sum of (i) and (ii) is the total effect of $X_a$ on $Y_a$

In case of the present study, outcome models and mediator models can be specified as follows:

Outcome models:

- $E[offspring's\ z - birth\ weight_a| \dots] = \beta_a GWG + \beta_m^T DNAm_a + \beta_c^T covariates_a$
- $E[offspring's\ z - birth\ weight_a| \dots] = \beta_a BMI + \beta_m^T DNAm_a + \beta_c^T covariates_a$

Mediator models:

- $E[DNAm_a| \dots] = \alpha_a GWG + \alpha_c covariates_a$
- $E[DNAm_a| \dots] = \alpha_a BMI + \alpha_c covariates_a$

(Where a ranges from 1 to N)

The total indirect (mediatory) effect $\alpha_a^T \beta_m$ and individual mediation contributions of a given $b$ mediator $(\alpha_a)b(\beta_m)b$ are estimated through a two-step process by the HDMA, HIMA, and HIMA2 methods, as stated below.

| Algorithm | R package | Description |
|---|---|---|
| HDMA | HDMED (Clark-Boucher et al, 2023) | Fits a high-dimensional mediation model with the de-biased LASSO approach as proposed by Guo et al. (2022), estimating the mediation contributions of potential mediators. The first step in HDMA is to perform sure independence screening (SIS) (Fan & Ly, 2008) to choose the "n_include" mediators that are most associated with the outcome (when Y is continuous) or the exposure (when Y is binary), based on *p*-values from linear regression. The second step is to fit the outcome model for the remaining mediators using de-sparsified (a.k.a. de-biased) LASSO, which as asymptotic properties allowing for computation of *p*-values by the "hdi" package. HDMA then fits the mediator models using linear regression among those mediators that have both survived SIS (in step 1) and been identified by the LASSO (in step 2), obtaining *p*-values for the mediation contributions by taking the maximum of the $\alpha_a$ and $\beta_m$ *p*-values. The global indirect effect is estimated by summing the mediation contributions, and the direct effect is estimated by subtracting the global indirect effect from an estimate of the total effect (Gao et al, 2019). |
| HIMA | HDMED (Clark-Boucher et al, 2023) | Fits a high-dimensional mediation model with the minimax concave penalty (MCP) as proposed by Zhang et al. (2016), estimating the mediation contributions of potential mediators. The first step in HIMA is to perform sure independence screening (SIS) to choose the "n_include" mediators that are most associated with the outcome (when Y is continuous) or the exposure (when Y is binary), based on p-values from linear regression. The second step is to apply the MCP to this subset of CpG sites for further dimension reduction. HIMA then fits the mediator models using linear regression among those mediators that have both survived SIS (in step 1) and been selected by the MCP (in step 2), which enables estimation of the mediation contributions. The global indirect effect is estimated by summing these contributions, and the direct effect is estimated by subtracting the global indirect effect from an estimate of the total effect. And *p*-values for the mediation contributions are computed by taking the maximum of the $\alpha_a$ and $\beta_m$ *p*-values, where the beta p-values are obtained via a second, unpenalized generalized linear model containing only the mediators selected by the MCP. |
| HIMA2 | HIMA (Zhang et al, 2016) | An upgraded version of HIMA for estimating and testing high-dimensional mediation effects. First, HIMA2 reduces the dimension of mediators to a manageable level based on the sure independence screening (SIS) method. Second, a de-biased Lasso procedure is implemented for further reducing dimensions and estimating regression parameters on the mediators that have survived both SIS (step 1) and de-biased LASSO (step 2). Third, a multiple-testing procedure is used to accurately control the false discovery rate (FDR) when testing high-dimensional mediation hypotheses (Perera et al, 2022) |

**Supplementary material 5: Directed acyclic graphs**

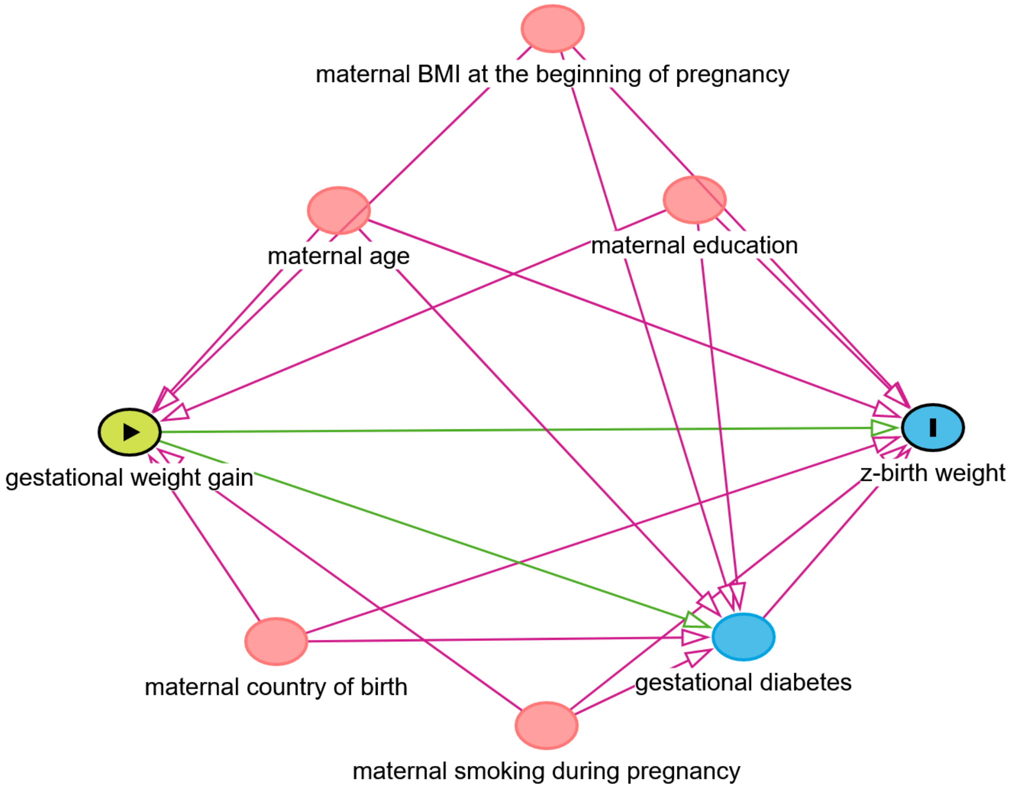


**Figure 1: Directed acyclic graph for the association between gestational weight gain and offspring's z-birth weight**

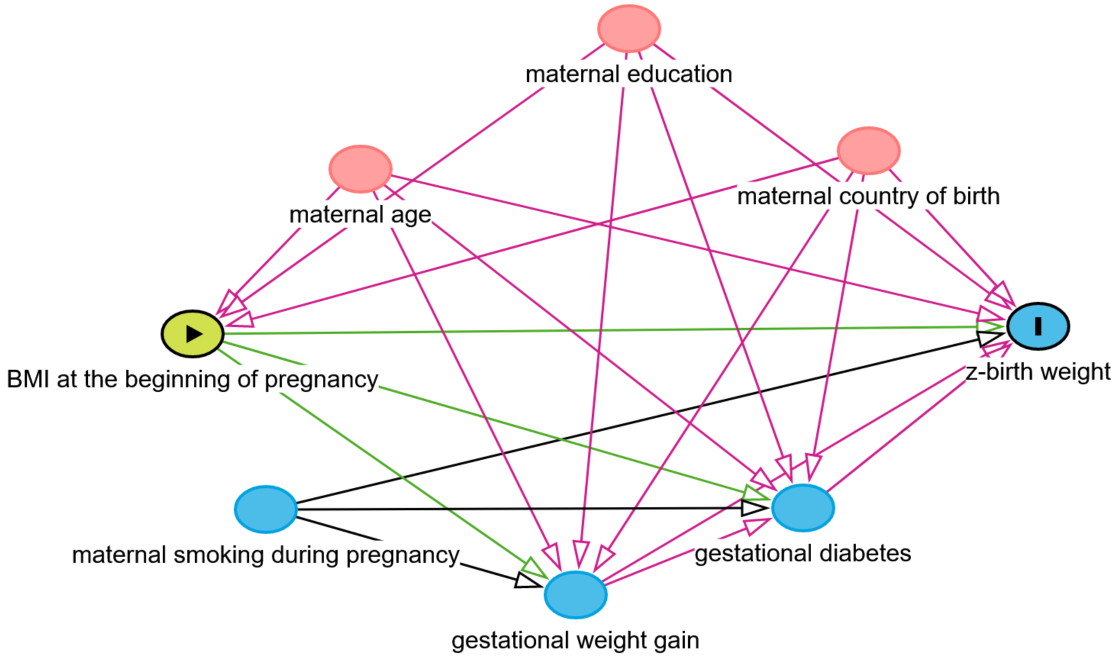


**Figure 2: Directed acyclic graph for the association of BMI at the beginning of pregnancy with offspring's z-birth weight**

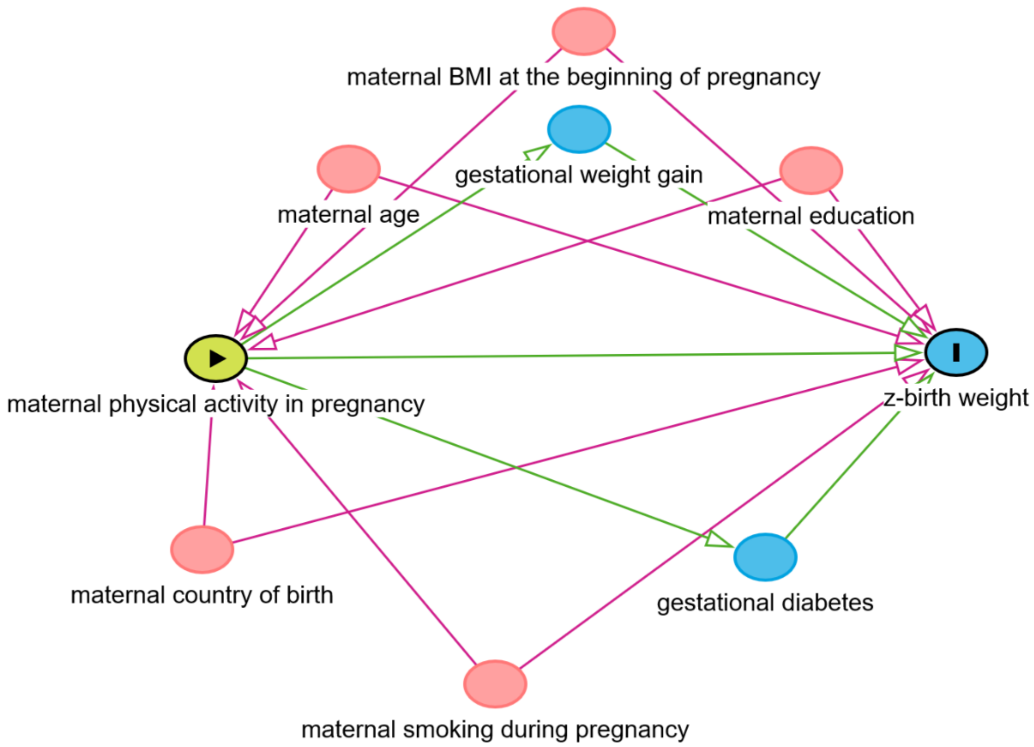


**Figure 3: Directed acyclic graph for the association between physical activity in pregnancy and offspring's z-birth weight**

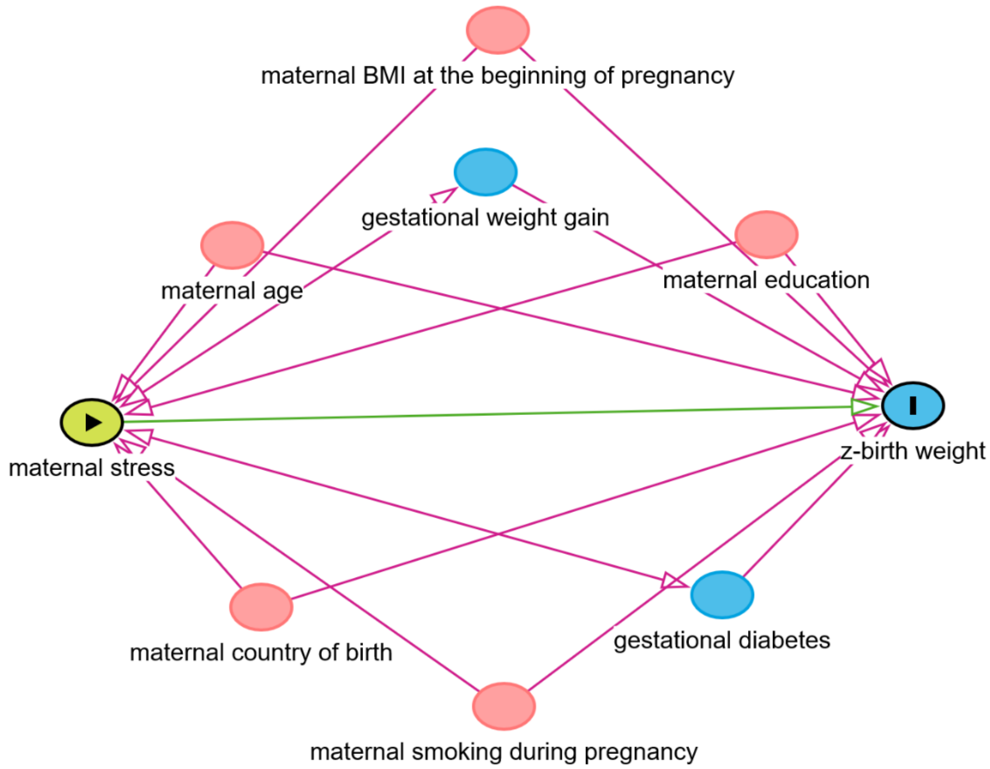


**Figure 4: Directed acyclic graph for the association between stress in pregnancy and offspring's z-birth weight**

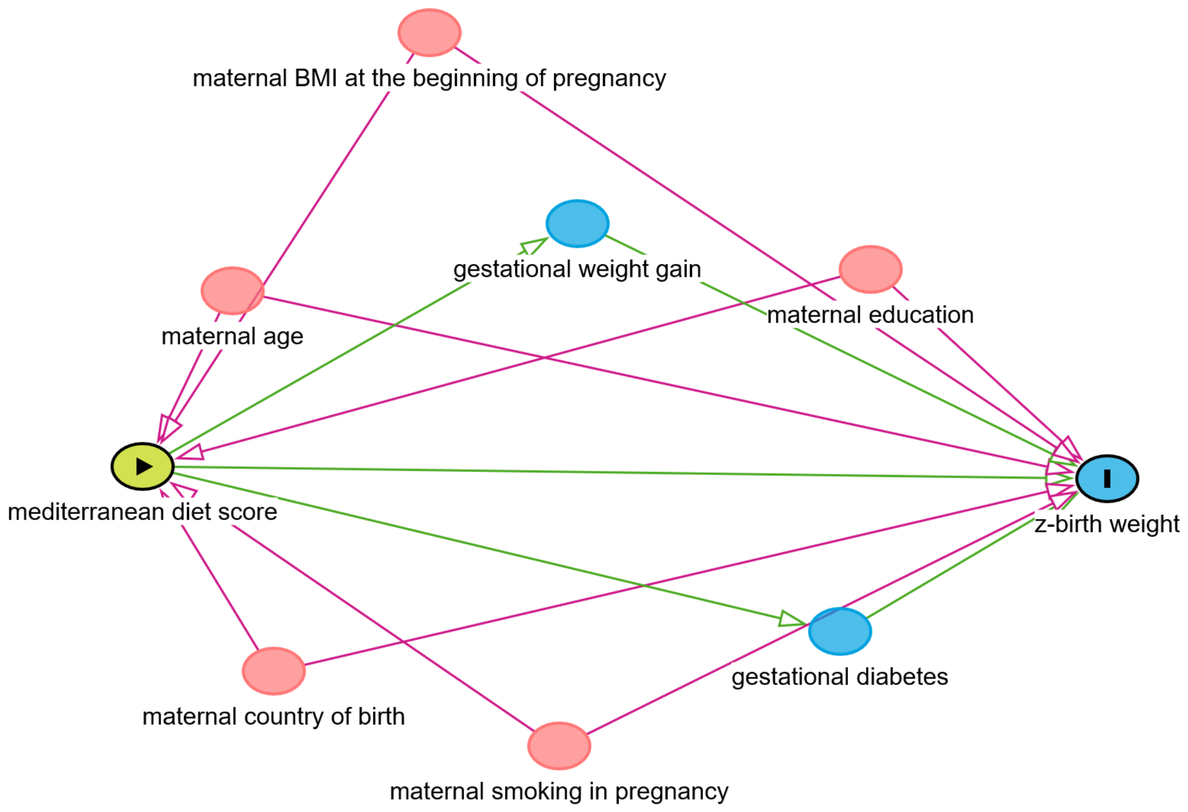


**Figure 5: Directed acyclic graph of the association between maternal mediterranean diet score and offspring's z-birth weight**

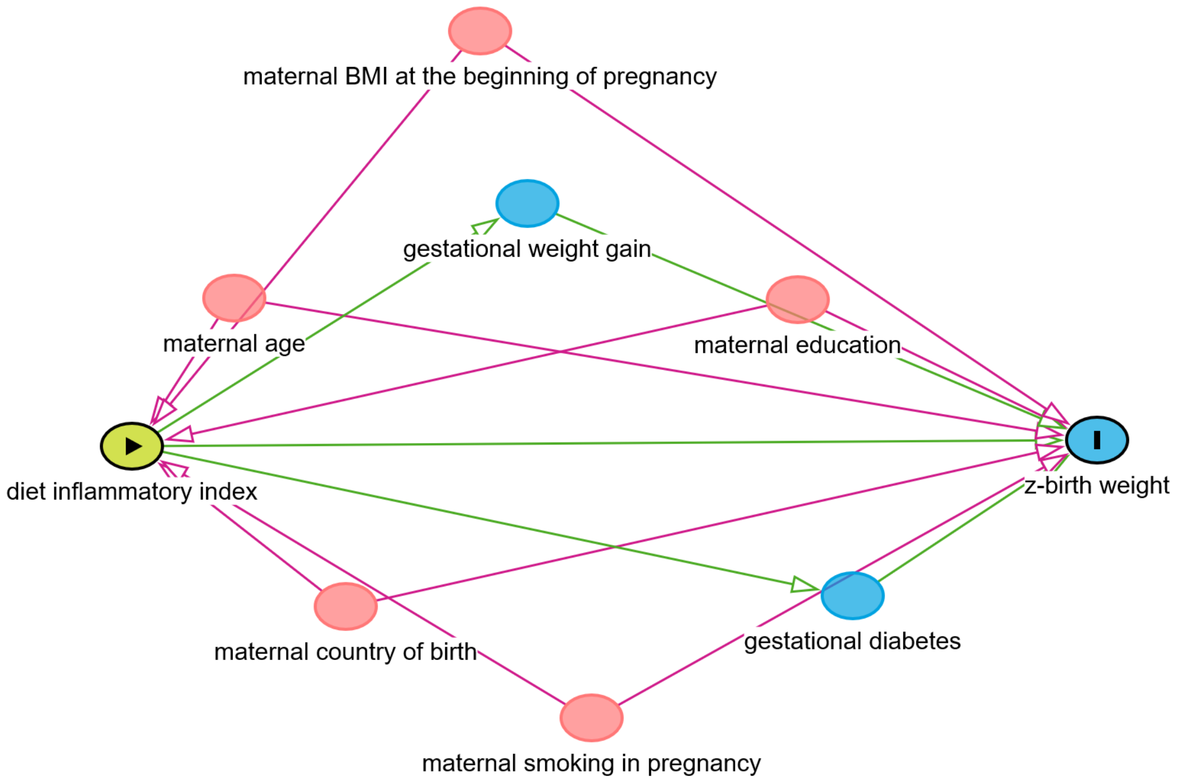


**Figure 6: Directed acyclic graph of the association between maternal diet inflammatory index and offspring's z-birth weight**

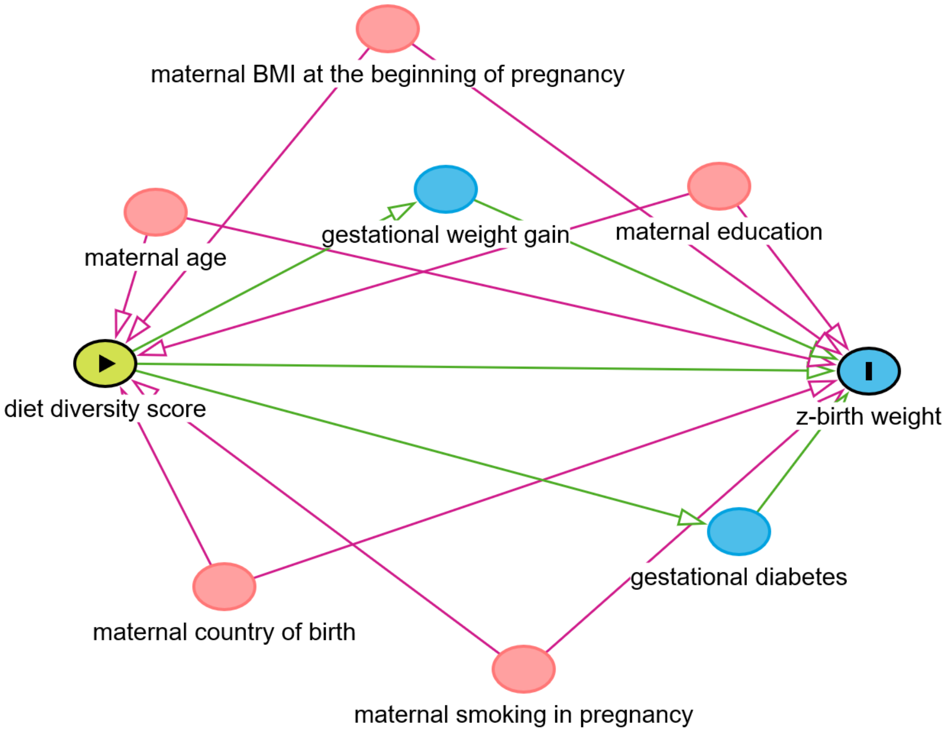


**Figure 7: Directed acyclic graph for the association between maternal diet diversity score and offspring's z-birth weight**

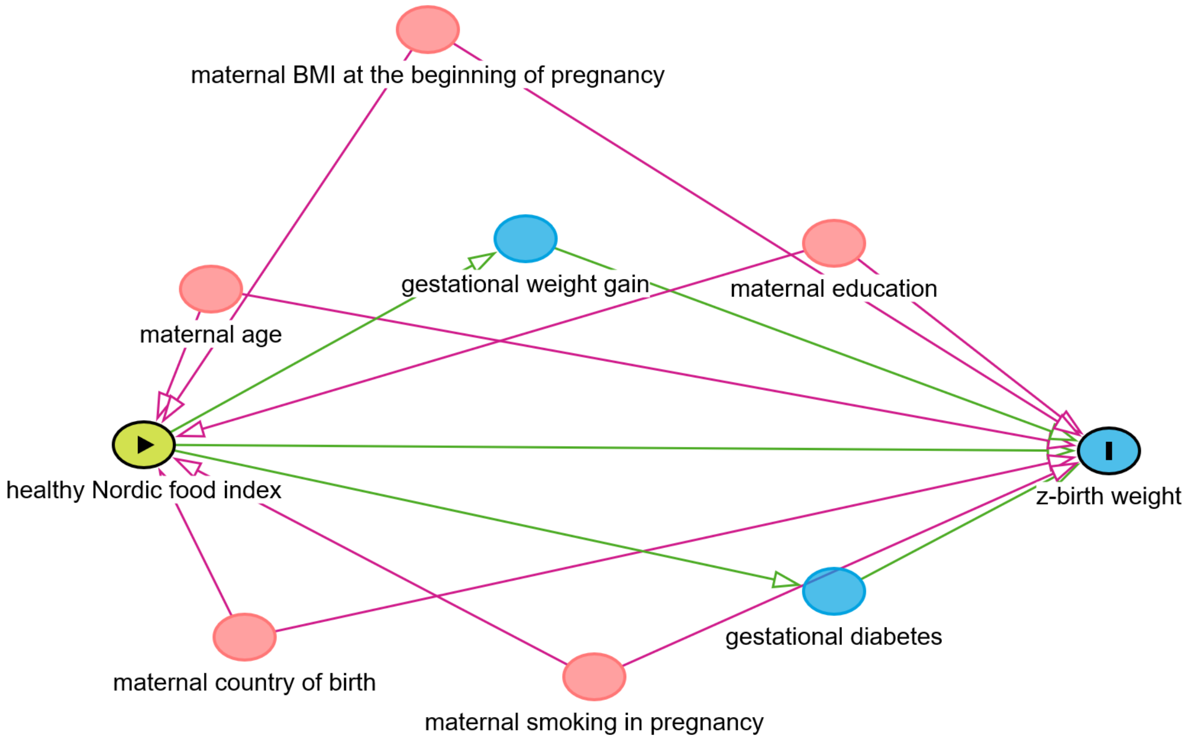


**Figure 8: Directed acyclic graph of the association between maternal healthy Nordic food index and offspring's z-birth weight**

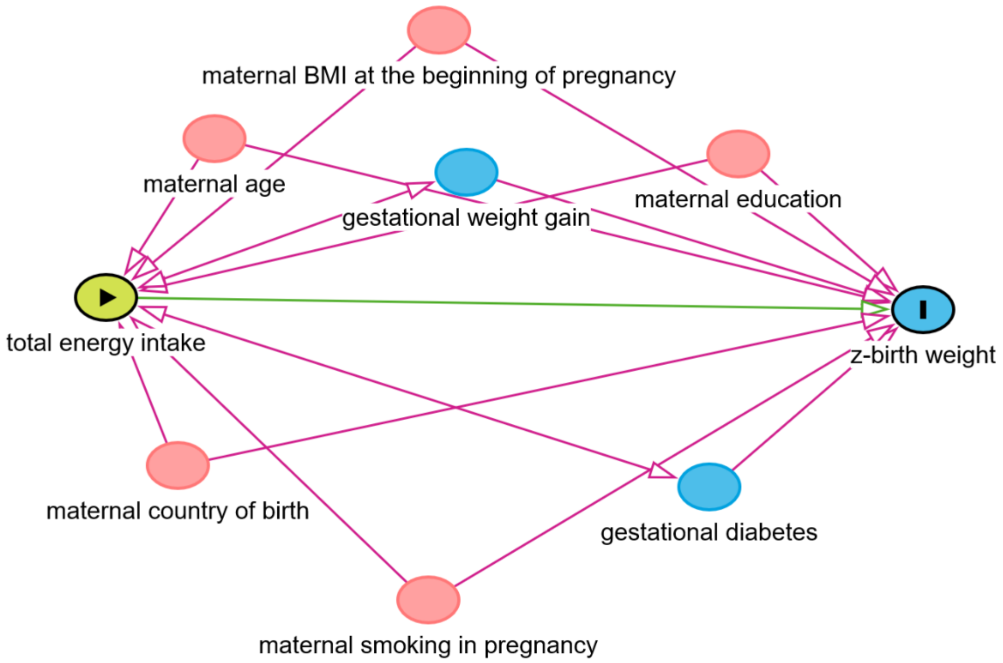


**Figure 9: Directed acyclic graph for the association between maternal total energy intake and offspring's z-birth weight**

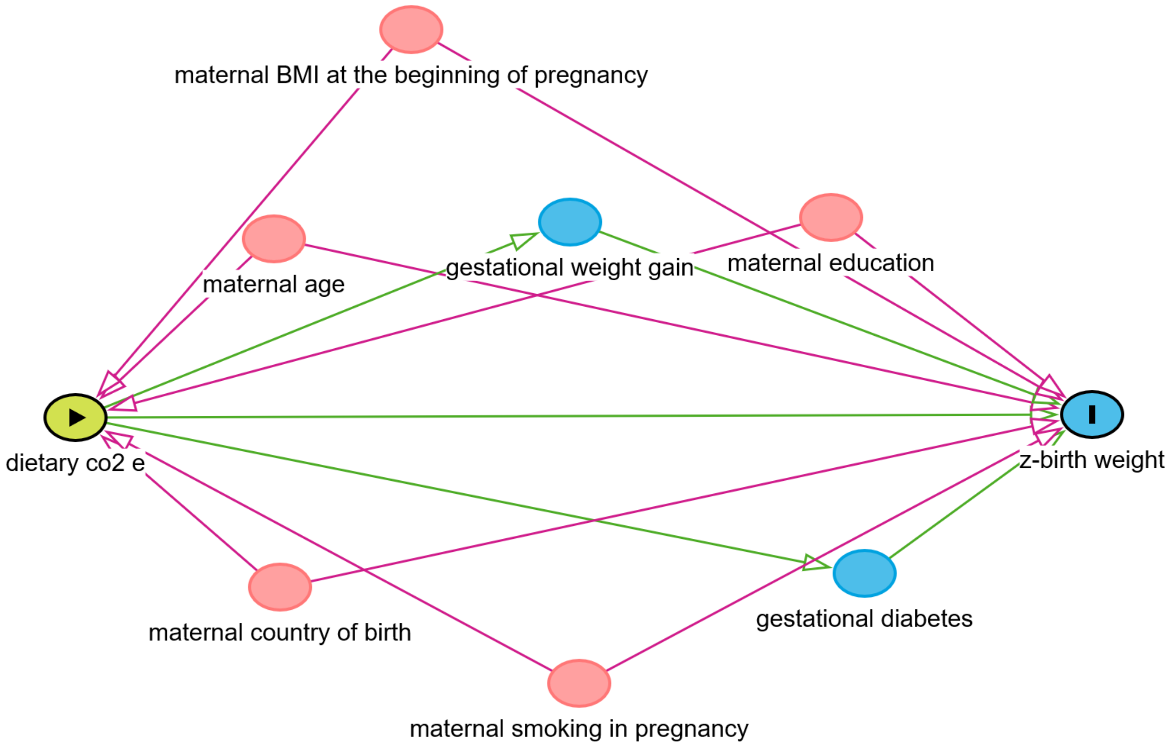


**Figure 10: Directed acyclic graph for the association between maternal dietary CO2 emissions/dietary carbon footprint and offspring's z-birth weight**

### Supplementary material 6: Details of the results of high dimensional mediation analysis

**Table 1: Details of the CpG sites selected as candidate mediators (n = 21) of the association between gestational weight gain and offspring's standardized birth weight by the HDMA approach**

| mediator | alpha | alpha_pv | beta | beta_pv | alpha*beta | ab_pv |
|---|---|---|---|---|---|---|
| cg04968127 | 1.2592562393181e-05 | 0.943482912347783 | 6.48304957559463 | 0.000472136630128327 | 8.16382062787609e-05 | 0.943482912347783 |
| **cg14798382** | **0.000179842458782697** | **0.00634143074595538** | **8.99872499684574** | **0.0492221696292345** | **0.00161835282934206** | **0.0492221696292345** |
| **cg21516291** | **0.000382824953097754** | **0.0583456464895654** | **3.05673551872342** | **0.0171201955417699** | **0.00117019463158753** | **0.0493456464895654** |
| cg27053299 | -9.51380933867518e-05 | 0.819666864942751 | -1.34195705859435 | 0.0366016746042471 | 0.00012767123596156 | 0.819666864942751 |
| cg10660916 | -0.000205810561889876 | 0.315750857812919 | 3.2975632784644 | 0.0127387091087569 | -0.000678673351208179 | 0.315750857812919 |
| cg14556683 | 3.12283125351938e-05 | 0.554971262719947 | 9.78276959624868 | 0.0311871443338365 | 0.000305499386411446 | 0.554971262719947 |
| cg10178960 | -1.94479367272413e-05 | 0.82559454162065 | -11.2477692170418 | 0.00598641837578967 | 0.000218745904055641 | 0.82559454162065 |
| cg16752400 | 0.000400552446129279 | 0.160401470771189 | 2.37681014163734 | 0.0159362145400034 | 0.000952037116217713 | 0.160401470771189 |
| cg09247736 | -2.28279102459041e-05 | 0.632406806124404 | 14.9861465912769 | 0.00252788271436388 | -0.000342102409317631 | 0.632406806124404 |
| cg18137450 | 8.61002142411789e-06 | 0.966710683317889 | 3.45867502348622 | 0.0130602143058213 | 2.97792660512778e-05 | 0.966710683317889 |
| cg07002832 | 0.000122424251695494 | 0.472365290755901 | -4.37769511791223 | 0.0145368026395345 | -0.000535936048961421 | 0.472365290755901 |
| cg02832224 | 0.000192465251885373 | 0.336575557147282 | 4.77028530035463 | 0.000838173914696004 | 0.000918114161897846 | 0.336575557147282 |
| cg05779272 | -8.55784564816783e-05 | 0.612011956298708 | 4.28096332247215 | 0.00899726103408218 | -0.000366358233391844 | 0.612011956298708 |
| **cg19242268** | **0.000760024753338449** | **0.00334890784466188** | **2.33829417241514** | **0.0167099222305364** | **0.00177716145162255** | **0.0167099222305364** |
| cg16402875 | 0.000116966565807042 | 0.438570007278646 | 4.33269171415938 | 0.0357029301538901 | 0.000506780070505849 | 0.438570007278646 |
| cg15672022 | 0.000155037752532813 | 0.714240985681105 | -1.98704121726876 | 0.0193976826517957 | -0.000308066404515413 | 0.714240985681105 |
| cg13131501 | 0.000318220164252777 | 0.130662136758009 | 3.43822256635646 | 0.012128154820652 | 0.00109411174980356 | 0.130662136758009 |
| cg04457572 | -0.00030989435514351 | 0.2017398152483 | -3.96635481787463 | 0.000496260108866941 | 0.00122915096855561 | 0.2017398152483 |
| cg01940139 | -2.51115958467901e-06 | 0.973494301559011 | 10.6946429024611 | 0.0171917815999335 | -2.68559550292346e-05 | 0.973494301559011 |
| **cg08461903** | **-0.000655509718183357** | **0.00811177347398761** | **-2.15793857112449** | **0.0372974015326812** | **0.00141454970461481** | **0.0372974015326812** |
| cg00154986 | -0.000238410807150942 | 0.329974294873344 | -3.39742352539049 | 0.00254938830678076 | 0.000809982484921947 | 0.329974294873344 |

alpha = coefficient estimate of the exposure (X) –> mediator (M) association; alpha_pv = p-value for the exposure (X) –> mediator (M) association; beta = coefficient estimate of the mediator (M) –> outcome (Y) association, adjusted for exposure; beta_pv = p-value for the mediator (M) –> outcome (Y) association, adjusted for exposure; alpha*beta = mediation effect; ab_pv = multiple-testing corrected p-value for statistical significance of the mediator

**Table 2: Details of the CpG sites selected as candidate mediators (n = 24) of the association between gestational weight gain and offspring's standardized birth weight by the HIMA approach**

| mediator | alpha | alpha_pv | beta | beta_pv | alpha_beta | ab_pv |
|---|---|---|---|---|---|---|
| cg04968127 | 1.2592562393181e-05 | 0.943482912347783 | 6.75958264553157 | 3.70754449405406e-05 | 8.51204662157196e-05 | 0.943482912347783 |
| cg05349624 | -4.35252966672858e-05 | 0.904077970813796 | 0.840322689378256 | 0.0578467395585855 | -3.657529435144e-05 | 0.904077970813796 |
| **cg21516291** | **0.000382824953097754** | **0.0583456464895654** | **1.69249274542587** | **0.0457505605120574** | **0.000647928455885948** | **0.0483456464895654** |
| cg27053299 | -9.51380933867518e-05 | 0.819666864942751 | -1.21501286390417 | 0.0418674077628547 | 0.000115594007312219 | 0.819666864942751 |
| cg10660916 | -0.000205810561889876 | 0.315750857812919 | 1.30803701933125 | 0.014709737663736 | -0.000269207833921323 | 0.315750857812919 |
| cg14556683 | 3.12283125351938e-05 | 0.554971262719947 | 3.85469332501059 | 0.0671005407551571 | 0.000120375567880756 | 0.554971262719947 |
| cg10178960 | -1.94479367272413e-05 | 0.82559454162065 | -9.80206704315084 | 5.62343191014168e-07 | 0.000190629979651374 | 0.82559454162065 |
| cg16752400 | 0.000400552446129279 | 0.160401470771189 | 0.928010378751901 | 0.0389988398009207 | 0.000371716827242432 | 0.160401470771189 |
| cg05304729 | 0.000478483687967837 | 0.156837160881313 | 1.20103752165605 | 0.0836401668609472 | 0.000574676862749736 | 0.156837160881313 |
| cg04751761 | -5.67505287452858e-05 | 0.807817547708683 | 0.814647372936708 | 0.143972412378901 | -4.62316691551162e-05 | 0.807817547708683 |
| cg09247736 | -2.28279102459041e-05 | 0.632406806124404 | 13.2141401690776 | 0.00184877684309049 | -0.000301651205756499 | 0.632406806124404 |
| cg18137450 | 8.61002142411789e-06 | 0.966710683317889 | 3.11429286252577 | 0.0173384770991019 | 2.68141282673243e-05 | 0.966710683317889 |
| cg02832224 | 0.000192465251885373 | 0.336575557147282 | 4.30528808756109 | 0.00114327472651364 | 0.000828618356211542 | 0.336575557147282 |
| cg05779272 | -8.55784564816783e-05 | 0.612011956298708 | 2.92931280928598 | 0.00379852288395121 | -0.000250686068770703 | 0.612011956298708 |

| | | | | | | |
|---|---|---|---|---|---|---|
| **cg19242268** | **0.000760024753338449** | **0.00334890784466188** | **1.3527834007674** | **0.0425573491051202** | **0.0010281488704886** | **0.0425573491051202** |
| cg05560494 | -0.000308318008237926 | 0.0449686663943078 | 1.07431757013899 | 0.111659504860774 | -0.000331231453440261 | 0.111659504860774 |
| cg16402875 | 0.000116966565807042 | 0.438570007278646 | 1.64915983572727 | 0.160556634901143 | 0.000192896562451925 | 0.438570007278646 |
| cg13131501 | 0.000318220164252777 | 0.130662136758009 | 3.05258313184316 | 0.00765833296821208 | 0.000971393505610388 | 0.130662136758009 |
| cg12804755 | -0.000991844089265759 | 0.0235692311108214 | -0.115673811519423 | 0.15428781303189 | 0.000114730386238381 | 0.15428781303189 |
| cg04457572 | -0.00030989435514351 | 0.2017398152483 | -4.40682206507038 | 0.000761053860591922 | 0.00136564928208718 | 0.2017398152483 |
| cg22247250 | -2.89695543774692e-05 | 0.904668336162854 | -0.207715197120164 | 0.261250052093978 | 6.01741669799932e-06 | 0.904668336162854 |
| **cg08461903** | **-0.000655509718183357** | **0.00811177347398761** | **-1.37189363084973** | **0.0577926151259687** | **0.000899289607335848** | **0.0477926151259687** |
| cg00154986 | -0.000238410807150942 | 0.329974294873344 | -3.19555112675531 | 0.00533293062193004 | 0.000761853923421836 | 0.329974294873344 |
| cg12145085 | 2.25872069728283e-05 | 0.803337553572515 | 3.78374200737788 | 0.0517847807552656 | 8.54641638524291e-05 | 0.803337553572515 |

alpha = coefficient estimate of the exposure (X) –> mediator (M) association; alpha_pv = p-value for the exposure (X) –> mediator (M) association; beta = coefficient estimate of the mediator (M) –> outcome (Y) association, adjusted for exposure; beta_pv = p-value for the mediator (M) –> outcome (Y) association, adjusted for exposure; alpha*beta = mediation effect; ab_pv = multiple-testing corrected p-value for statistical significance of the mediator

**Table 3: Details of the CpG sites selected as candidate mediators (n = 24) of the association between gestational weight gain and offspring's standardized birth weight by the HIMA2 approach**

| probe | alpha | beta | alpha*beta | BH.FDR |
|---|---|---|---|---|
| cg04968127 | 1.2592562393181e-05 | 6.75958264553157 | 8.51204662157196e-05 | 0.966710683317889 |
| cg05349624 | -4.35252966672858e-05 | 0.840322689378256 | -3.657529435144e-05 | 0.966710683317889 |
| cg21516291 | 0.000382824953097754 | 1.69249274542587 | 0.000647928455885948 | 0.280059103149914 |
| cg27053299 | -9.51380933867518e-05 | -1.21501286390417 | 0.000115594007312219 | 0.966710683317889 |
| cg10660916 | -0.000205810561889876 | 1.30803701933125 | -0.000269207833921323 | 0.673151114294564 |
| cg14556683 | 3.12283125351938e-05 | 3.85469332501059 | 0.000120375567880756 | 0.948610209186606 |
| cg10178960 | -1.94479367272413e-05 | -9.80206704315084 | 0.000190629979651374 | 0.966710683317889 |
| cg16752400 | 0.000400552446129279 | 0.928010378751901 | 0.000371716827242432 | 0.481204412313568 |
| cg05304729 | 0.000478483687967837 | 1.20103752165605 | 0.000574676862749736 | 0.481204412313568 |
| cg04751761 | -5.67505287452858e-05 | 0.814647372936708 | -4.62316691551162e-05 | 0.966710683317889 |
| cg09247736 | -2.28279102459041e-05 | 13.2141401690776 | -0.000301651205756499 | 0.948610209186606 |
| cg18137450 | 8.61002142411789e-06 | 3.11429286252577 | 2.68141282673243e-05 | 0.966710683317889 |
| cg02832224 | 0.000192465251885373 | 4.30528808756109 | 0.000828618356211542 | 0.673151114294564 |
| cg05779272 | -8.55784564816783e-05 | 2.92931280928598 | -0.000250686068770703 | 0.948610209186606 |
| **cg19242268** | **0.000760024753338449** | **1.3527834007674** | **0.0010281488704886** | **0.049037378827188** |
| cg05560494 | -0.000308318008237926 | 1.07431757013899 | -0.000331231453440261 | 0.269811998365847 |
| cg16402875 | 0.000116966565807042 | 1.64915983572727 | 0.000192896562451925 | 0.809667705745192 |
| cg13131501 | 0.000318220164252777 | 3.05258313184316 | 0.000971393505610388 | 0.481204412313568 |
| cg12804755 | -0.000991844089265759 | -0.115673811519423 | 0.000114730386238381 | 0.188553848886571 |
| cg04457572 | -0.00030989435514351 | -4.40682206507038 | 0.00136564928208718 | 0.537972840662133 |
| cg22247250 | -2.89695543774692e-05 | -0.207715197120164 | 6.01741669799932e-06 | 0.966710683317889 |
| **cg08461903** | **-0.000655509718183357** | **-1.37189363084973** | **0.000899289607335848** | **0.049734128168785** |
| cg00154986 | -0.000238410807150942 | -3.19555112675531 | 0.000761853923421836 | 0.673151114294564 |
| cg12145085 | 2.25872069728283e-05 | 3.78374200737788 | 8.54641638524291e-05 | 0.966710683317889 |

alpha = coefficient estimates of exposure (X) –> mediators (M) associations; beta = coefficient estimates of mediators (M) –> outcome (Y) associations, adjusted for exposure; alpha*beta = mediation effects; BH.FDR = Benjamini-Hochberg FDR adjusted p-value for statistical significance of the mediator

**Table 4: Details of the CpG sites selected as candidate mediators (n = 25) of the association between maternal BMI at the beginning of pregnancy and offspring's standardized birth weight by the HDMA approach**

| mediator | alpha | alpha_pv | beta | beta_pv | alpha*beta | ab_pv |
|---|---|---|---|---|---|---|
| cg04968127 | -0.000117134888335002 | 0.618917142513759 | 5.27195794579884 | 0.00413647459914182 | -0.000617530205287973 | 0.618917142513759 |
| cg21516291 | 0.00033374992013122 | 0.21164768787333 | 2.59878056884976 | 0.0358030566475134 | 0.000867342807292175 | 0.21164768787333 |
| cg23260105 | 2.90524138089407e-05 | 0.51073316100907 | -22.7082985275131 | 0.00388630875461215 | -0.000659730885718269 | 0.51073316100907 |
| cg00376553 | -0.000320823988220308 | 0.489055912097046 | 1.57548277360594 | 0.0480063334806574 | -0.000505452666800651 | 0.489055912097046 |
| **cg17040807** | **-0.000704476905025845** | **0.0159508530025362** | **-3.07893622985714** | **0.0111265033324041** | **0.0021690394659817** | **0.0159508530025362** |
| cg21649604 | -2.17818339879823e-05 | 0.939581490118541 | 2.80143500194308 | 0.0481309517778563 | -6.10203921404471e-05 | 0.939581490118541 |
| cg16752400 | 0.000656226879343457 | 0.080350059076816 | 3.71635251500328 | 0.000128013654566387 | 0.00243877041346081 | 0.080350059076816 |
| cg25494075 | -3.91488935606756e-05 | 0.825774942846241 | 4.90109031266213 | 0.0409676393419151 | -0.000191872262981668 | 0.825774942846241 |
| cg08289567 | 0.000275383318912381 | 0.0981604838802426 | -4.71732003165868 | 0.027716787783651 | -0.00129907124669003 | 0.0981604838802426 |
| cg18137450 | 6.37984648871824e-05 | 0.813541141875918 | 5.25562586924733 | 0.000130841314467748 | 0.000335300862479344 | 0.813541141875918 |
| cg02832224 | 0.000204165898093526 | 0.439698016260347 | 5.05210158320132 | 0.00027709340581825 | 0.00103146685699402 | 0.439698016260347 |
| cg05779272 | -1.89362646499239e-05 | 0.932030953822198 | 3.25181215955961 | 0.0383588085887173 | -6.15771756452615e-05 | 0.932030953822198 |
| **cg19242268** | **0.000908375085696785** | **0.00839714356148382** | **2.58860418310286** | **0.00588795295016003** | **0.00235142354666111** | **0.00839714356148382** |
| **cg26552621** | **0.000850935104261077** | **0.0139543437332839** | **-2.94899820527688** | **0.0173224491001761** | **-0.00250940609527301** | **0.0173224491001761** |
| cg18034719 | 2.94817270353869e-05 | 0.883337290484565 | 6.58689405871855 | 0.000137747152649459 | 0.000194193012650152 | 0.883337290484565 |
| cg09171931 | 0.000195477624652817 | 0.231865982738286 | 4.51454056547862 | 0.0488914826091835 | 0.000882491666138544 | 0.231865982738286 |
| cg13131501 | -4.76366283295481e-05 | 0.863672121362842 | 3.52422373071971 | 0.00970500342750538 | -0.000167882136010468 | 0.863672121362842 |
| cg03688987 | -0.000302895818199109 | 0.329872638719241 | -2.57539772364163 | 0.0199558712340388 | 0.000780077200690554 | 0.329872638719241 |
| **cg04457572** | **-0.000694880116204726** | **0.031228407402061** | **-2.95153383564912** | **0.00837106340936899** | **0.00205096217469804** | **0.031228407402061** |
| **cg06457011** | **0.00177574711976154** | **0.00846277004263347** | **1.10692179627811** | **0.0475767762890234** | **0.00196561319154213** | **0.0475767762890234** |
| cg14787880 | -1.59799882978612e-05 | 0.82367882718336 | 12.4236595667117 | 0.00412635467076922 | -0.000198529934492664 | 0.82367882718336 |
| cg01940139 | 2.37574795826187e-05 | 0.812076779671485 | 10.6467896073137 | 0.0137169578521857 | 0.000252940886716191 | 0.812076779671485 |
| cg08461903 | -0.000265659351759233 | 0.41928543011665 | -2.04957775316619 | 0.0418100945645003 | 0.000544489497286276 | 0.41928543011665 |
| cg00154986 | -0.000431642782876597 | 0.183246804486515 | -2.42207986145519 | 0.0250716193415645 | 0.00104547329174788 | 0.183246804486515 |
| cg12145085 | 0.000114330079463317 | 0.341439491945482 | 7.07713886503554 | 0.0311642639360312 | 0.000809129848812443 | 0.341439491945482 |

alpha = coefficient estimate of the exposure (X) –> mediator (M) association; alpha_pv = p-value for the exposure (X) –> mediator (M) association; beta = coefficient estimate of the mediator (M) –> outcome (Y) association, adjusted for exposure; beta_pv = p-value for the mediator (M) –> outcome (Y) association, adjusted for exposure; alpha*beta = mediation effect; ab_pv = multiple-testing corrected p-value for statistical significance of the mediator

**Table 5: Details of the CpG sites selected as candidate mediators (n = 24) of the association between maternal BMI at the beginning of pregnancy and offspring's standardized birth weight by the HIMA approach**

| mediator | alpha | alpha_pv | beta | beta_pv | alpha*beta | ab_pv |
|---|---|---|---|---|---|---|
| cg04968127 | -0.000117134888335002 | 0.618917142513759 | 5.37173490299259 | 0.000290892720913403 | -0.000629217568027269 | 0.618917142513759 |
| cg14798382 | 0.000162532316756727 | 0.0591352611892092 | 3.90604522473127 | 0.0696595668639727 | 0.000634858579732123 | 0.0696595668639727 |
| cg21516291 | 0.00033374992013122 | 0.21164768787333 | 1.26334937860766 | 0.0177476188496189 | 0.000421642754208133 | 0.21164768787333 |
| cg23260105 | 2.90524138089407e-05 | 0.51073316100907 | -21.342802189629 | 0.00193441981218922 | -0.000620059921055468 | 0.51073316100907 |
| cg00376553 | -0.000320823988220308 | 0.489055912097046 | 2.04225736532118 | 0.00268698953410555 | -0.000655205152914639 | 0.489055912097046 |
| **cg17040807** | **-0.000704476905025845** | **0.0159508530025362** | **-1.53143545695465** | **0.00660634221675107** | **0.00107886091096226** | **0.0159508530025362** |
| cg16752400 | 0.000656226879343457 | 0.080350059076816 | 2.68237575728844 | 0.000417339650600328 | 0.00176024707243193 | 0.080350059076816 |
| cg05304729 | 0.000734327829433121 | 0.0967000500853513 | 1.37499408005475 | 0.0185295353501625 | 0.00100969641828999 | 0.0967000500853513 |
| cg15482893 | 0.000823698172114162 | 0.0788628214820748 | -0.0941678797143056 | 0.00685939995463376 | -7.75659103925398e-05 | 0.0788628214820748 |
| cg18137450 | 6.37984648871824e-05 | 0.813541141875918 | 4.66617476087281 | 3.43971182027634e-05 | 0.000297694786639001 | 0.813541141875918 |
| cg02832224 | 0.000204165898093526 | 0.439698016260347 | 4.71846067911673 | 3.31097297124896e-05 | 0.000963348762170857 | 0.439698016260347 |
| cg05779272 | -1.89362646499239e-05 | 0.932030953822198 | 0.729871283409996 | 0.0409377826375437 | -1.38210357830313e-05 | 0.932030953822198 |

| | | | | | | |
|---|---|---|---|---|---|---|
| **cg19242268** | **0.000908375085696785** | **0.00839714356148382** | **2.65827072473442** | **0.0026266133470941** | **0.00241470689738589** | **0.00839714356148382** |
| **cg26552621** | **0.000850935104261077** | **0.0139543437332839** | **-3.62251245003404** | **5.0292368803488e-05** | **-0.00308252300935677** | **0.0139543437332839** |
| cg18034719 | 2.94817270353869e-05 | 0.883337290484565 | 7.11023506285039 | 1.22203072819428e-05 | 0.000209622009280392 | 0.883337290484565 |
| cg13131501 | -4.76366283295481e-05 | 0.863672121362842 | 2.4167626589876 | 0.00145077902020376 | -0.000115126424546923 | 0.863672121362842 |
| cg03688987 | -0.000302895818199109 | 0.329872638719241 | -3.20384719678319 | 0.00292620695547031 | 0.000970431918054567 | 0.329872638719241 |
| **cg04457572** | **-0.000694880116204726** | **0.031228407402061** | **-1.87910302900048** | **0.0115078360745678** | **0.00130575133115251** | **0.031228407402061** |
| **cg06457011** | **0.00177574711976154** | **0.00846277004263347** | **0.278375863358203** | **0.0321060839835381** | **0.000494325137569461** | **0.0321060839835381** |
| cg14787880 | -1.59799882978612e-05 | 0.82367882718336 | 13.5024815599172 | 0.00108207855642818 | -0.000215769497319563 | 0.82367882718336 |
| cg01940139 | 2.37574795826187e-05 | 0.812076779671485 | 7.9930068442057 | 0.00268393072307276 | 0.000189893696904948 | 0.812076779671485 |
| cg08461903 | -0.000265659351759233 | 0.41928543011665 | -2.26673374873755 | 0.0111202593175418 | 0.000602179018300393 | 0.41928543011665 |
| cg00154986 | -0.000431642782876597 | 0.183246804486515 | -0.321149817365529 | 0.0802809621460652 | 0.000138622000887968 | 0.183246804486515 |
| cg05632420 | 0.000366182079550837 | 0.313439226072183 | -1.54941738008431 | 0.00164985898614122 | -0.000567368878331482 | 0.313439226072183 |

alpha = coefficient estimate of the exposure (X) –> mediator (M) association; alpha_pv = p-value for the exposure (X) –> mediator (M) association; beta = coefficient estimate of the mediator (M) –> outcome (Y) association, adjusted for exposure; beta_pv = p-value for the mediator (M) –> outcome (Y) association, adjusted for exposure; alpha*beta = mediation effect; ab_pv = multiple-testing corrected p-value for statistical significance of the mediator

**Table 6: Details of the CpG sites selected as candidate mediators (n = 24) of the association between maternal BMI at the beginning of pregnancy and offspring's standardized birth weight by the HIMA2 approach**

| **mediator** | **alpha** | **beta** | **alpha*beta** | **BH.FDR** |
|---|---|---|---|---|
| cg04968127 | -0.000117134888335002 | 5.37173490299259 | -0.000629217568027269 | 0.825222856685011 |
| cg14798382 | 0.000162532316756727 | 3.90604522473127 | 0.000634858579732123 | 0.236541044756837 |
| cg21516291 | 0.00033374992013122 | 1.26334937860766 | 0.000421642754208133 | 0.461776773541811 |
| cg23260105 | 2.90524138089407e-05 | -21.342802189629 | -0.000620059921055468 | 0.721035050836334 |
| cg00376553 | -0.000320823988220308 | 2.04225736532118 | -0.000655205152914639 | 0.721035050836334 |
| cg17040807 | -0.000704476905025845 | -1.53143545695465 | 0.00107886091096226 | 0.0957051180152175 |
| cg16752400 | 0.000656226879343457 | 2.68237575728844 | 0.00176024707243193 | 0.241050177230448 |
| cg05304729 | 0.000734327829433121 | 1.37499408005475 | 0.00100969641828999 | 0.257866800227604 |
| cg15482893 | 0.000823698172114162 | -0.0941678797143056 | -7.75659103925398e-05 | 0.241050177230448 |
| cg18137450 | 6.37984648871824e-05 | 4.66617476087281 | 0.000297694786639001 | 0.921743259636068 |
| cg02832224 | 0.000204165898093526 | 4.71846067911673 | 0.000963348762170857 | 0.703516826016555 |
| cg05779272 | -1.89362646499239e-05 | 0.729871283409996 | -1.38210357830313e-05 | 0.932030953822198 |
| cg19242268 | 0.000908375085696785 | 2.65827072473442 | 0.00241470689738589 | 0.0957051180152175 |
| cg26552621 | 0.000850935104261077 | -3.62251245003404 | -0.00308252300935677 | 0.0957051180152175 |
| cg18034719 | 2.94817270353869e-05 | 7.11023506285039 | 0.000209622009280392 | 0.921743259636068 |
| cg13131501 | -4.76366283295481e-05 | 2.4167626589876 | -0.000115126424546923 | 0.921743259636068 |
| cg03688987 | -0.000302895818199109 | -3.20384719678319 | 0.000970431918054567 | 0.608995640712446 |
| cg04457572 | -0.000694880116204726 | -1.87910302900048 | 0.00130575133115251 | 0.149896355529893 |
| cg06457011 | 0.00177574711976154 | 0.278375863358203 | 0.000494325137569461 | 0.0957051180152175 |
| cg14787880 | -1.59799882978612e-05 | 13.5024815599172 | -0.000215769497319563 | 0.921743259636068 |
| cg01940139 | 2.37574795826187e-05 | 7.9930068442057 | 0.000189893696904948 | 0.921743259636068 |
| cg08461903 | -0.000265659351759233 | -2.26673374873755 | 0.000602179018300393 | 0.703516826016555 |
| cg00154986 | -0.000431642782876597 | -0.321149817365529 | 0.000138622000887968 | 0.439792330767635 |
| cg05632420 | 0.000366182079550837 | -1.54941738008431 | -0.000567368878331482 | 0.608995640712446 |

alpha = coefficient estimates of exposure (X) –> mediators (M) associations; beta = coefficient estimates of mediators (M) –> outcome (Y) associations, adjusted for exposure; alpha*beta = mediation effects; BH.FDR = Benjamini-Hochberg FDR adjusted p-value for statistical significance of the mediator

### Supplementary material 7: Summary of results from ordinary least squares bootstrap mediation analysis with candidate CpG sites selected by high dimensional mediation analysis

| **Epigenetic mediation of the association between gestational weight gain and offspring's standardized birth weight** | | | | | | | | |
|---|---|---|---|---|---|---|---|---|
| **Single mediator models – OLS bootstrap approach** | | | | | | | | |
| **CpG** | **Indirect effect** | | **Direct effect** | | | **Total effect** | | |
| | **Estimate** | **95% CI** | **Estimate** | **SE** | **p-value** | **Estimate** | **SE** | **p-value** |
| **cg19242268** | 0.0035 | 0.0011, 0.0077 | 0.0269 | 0.0070 | 0.0001 | 0.0304 | 0.0071 | 0.00002 |
| **cg08461903** | 0.0030 | 0.0006, 0.0063 | 0.0274 | 0.0071 | 0.0001 | 0.0304 | 0.0071 | 0.00002 |
| **cg14798382** | 0.0042 | 0.0014, 0.0081 | 0.0262 | 0.0070 | 0.0001 | 0.0304 | 0.0071 | 0.00002 |
| cg21516291 | 0.0028 | -0.0001, 0.0065 | 0.0276 | 0.0068 | 0.00005 | 0.0304 | 0.0071 | 0.00002 |
| **Serial multiple mediator model – OLS bootstrap approach** | | | | | | | | |
| **Indirect effects** | | | **Direct effect** | | | **Total effect** | | |
| **Pathway** | **Estimate** | **95% CI** | **Estimate** | **SE** | **p-value** | **Estimate** | **SE** | **p-value** |
| **Total indirect** | 0.009345 | 0.005113, 0.0148 | 0.021091 | 0.006828 | 0.002 | 0.030436 | 0.007083 | 0.00002 |
| **Indirect1** | 0.003237 | 0.000947, 0.007157 | **(Indirect1: GWG -> cg19242268 -> z-birth weight)** | | | | | |
| **Indirect2** | 0.002556 | 0.000487, 0.005649 | **(Indirect2: GWG -> cg08461903 -> z-birth weight)** | | | | | |
| **Indirect3** | 0.002875 | 0.000567, 0.006130 | **(Indirect3: GWG -> cg14798382 -> z-birth weight)** | | | | | |
| Indirect4 | 0.000033 | -0.000203, 0.000295 | (Indirect4: GWG -> cg19242268 -> cg08461903 -> z-birth weight) | | | | | |
| **Indirect5** | 0.000269 | 0.000028, 0.000840 | **(Indirect5: GWG -> cg19242268 -> cg14798382 -> z-birth weight)** | | | | | |
| **Indirect6** | 0.000370 | 0.000068, 0.000986 | **(Indirect6: GWG -> cg08461903 -> cg14798382 -> z-birth weight)** | | | | | |
| Indirect7 | 0.000005 | -0.000026, 0.000055 | (Indirect7: GWG -> cg19242268 -> cg08461903 -> cg14798382 -> z-birth weight) | | | | | |
| **Parallel multiple mediator model – OLS bootstrap approach** | | | | | | | | |
| **Indirect effects** | | | **Direct effect** | | | **Total effect** | | |
| **Pathway** | **Estimate** | **95% CI** | **Estimate** | **SE** | **p-value** | **Estimate** | **SE** | **p-value** |
| **Total indirect** | 0.0093 | 0.0051, 0.0148 | 0.0211 | 0.0068 | 0.002 | 0.0304 | 0.0071 | 0.00002 |
| **cg19242268** | 0.0032 | 0.0009, 0.0071 | | | | | | |
| **cg08461903** | 0.0026 | 0.0005, 0.0057 | | | | | | |
| **cg14798382** | 0.0035 | 0.0012, 0.0070 | | | | | | |
| **Epigenetic mediation of the association between BMI at the beginning of pregnancy and offspring's standardized birth weight** | | | | | | | | |
| **Single mediator models – OLS bootstrap approach** | | | | | | | | |
| **CpG** | **Indirect effect** | | **Direct effect** | | | **Total effect** | | |
| | **Estimate** | **95% CI** | **Estimate** | **SE** | **p-value** | **Estimate** | **SE** | **p-value** |
| **cg17040807** | 0.0040 | 0.0010, 0.0087 | 0.0324 | 0.0095 | 0.0006 | 0.0364 | 0.0095 | 0.0001 |
| **cg19242268** | 0.0054 | 0.0017, 0.0109 | 0.0310 | 0.0093 | 0.0008 | 0.0364 | 0.0095 | 0.0001 |
| **cg26552621** | 0.0051 | 0.0011, 0.0105 | 0.0313 | 0.0096 | 0.001 | 0.0364 | 0.0095 | 0.0001 |
| **cg04457572** | 0.0052 | 0.0015, 0.0106 | 0.0312 | 0.0094 | 0.0009 | 0.0364 | 0.0095 | 0.0001 |
| **cg06457011** | 0.0041 | 0.0006, 0.0097 | 0.0323 | 0.0094 | 0.0006 | 0.0364 | 0.0095 | 0.0001 |
| **Serial multiple mediator model – OLS bootstrap approach** | | | | | | | | |
| **Indirect effects** | | | **Direct effect** | | | **Total effect** | | |
| **Pathway** | **Estimate** | **95% CI** | **Estimate** | **SE** | **p-value** | **Estimate** | **SE** | **p-value** |
| **Total indirect** | 0.01170 | 0.00567, 0.02007 | 0.02465 | 0.00935 | 0.008 | 0.03635 | 0.00947 | 0.0001 |
| **Indirect1** | 0.00455 | 0.00146, 0.00959 | **(Indirect1: BMI -> cg19242268 -> z-birth weight)** | | | | | |
| **Indirect2** | 0.00222 | 0.00021, 0.00605 | **(Indirect2: BMI -> cg26552621 -> z-birth weight)** | | | | | |
| **Indirect3** | 0.00308 | 0.00020, 0.00695 | **(Indirect3: BMI -> cg04457572 -> z-birth weight)** | | | | | |
| **Indirect4** | 0.00058 | 0.00013, 0.00161 | **(Indirect4: BMI -> cg19242268 -> cg26552621 -> z-birth weight)** | | | | | |
| Indirect5 | -0.00004 | -0.00051, 0.00031 | (Indirect5: BMI -> cg19242268 -> cg04457572 -> z-birth weight) | | | | | |
| **Indirect6** | 0.00104 | 0.00013, 0.00244 | **(Indirect6: BMI -> cg26552621 -> cg04457572 -> z-birth weight)** | | | | | |
| **Indirect7** | 0.00027 | 0.00008, 0.00067 | **(Indirect7: BMI -> cg19242268 -> cg26552621 -> cg04457572 -> z-birth weight)** | | | | | |
| **Parallel multiple mediator model – OLS bootstrap approach** | | | | | | | | |
| **Indirect effects** | | | **Direct effect** | | | **Total effect** | | |
| **Pathway** | **Estimate** | **95% CI** | **Estimate** | **SE** | **p-value** | **Estimate** | **SE** | **p-value** |
| **Total indirect** | 0.0136 | 0.0069, 0.0224 | 0.0228 | 0.0093 | 0.01 | 0.0364 | 0.0095 | 0.0001 |
| cg17040807 | 0.0017 | -0.00001, 0.0050 | | | | | | |
| **cg19242268** | 0.0045 | 0.0014, 0.0093 | | | | | | |
| cg26552621 | 0.0016 | -0.0002, 0.0052 | | | | | | |
| **cg04457572** | 0.0036 | 0.0009, 0.0081 | | | | | | |
| **cg06457011** | 0.0022 | 0.0002, 0.0064 | | | | | | |

**Supplementary material 8: Single- and multiple- mediator models (graphical presentations)**

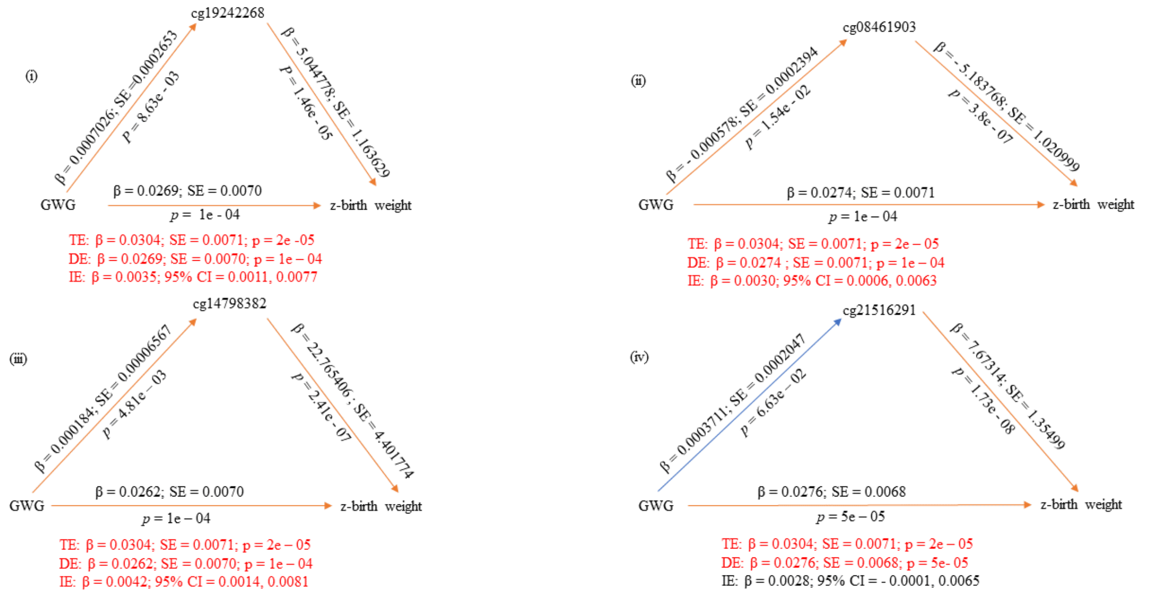


**Figure 1: Single mediator models as per the OLS bootstrap approach with candidate CpG sites as mediators of the association between GWG and offspring's z-birth weight**

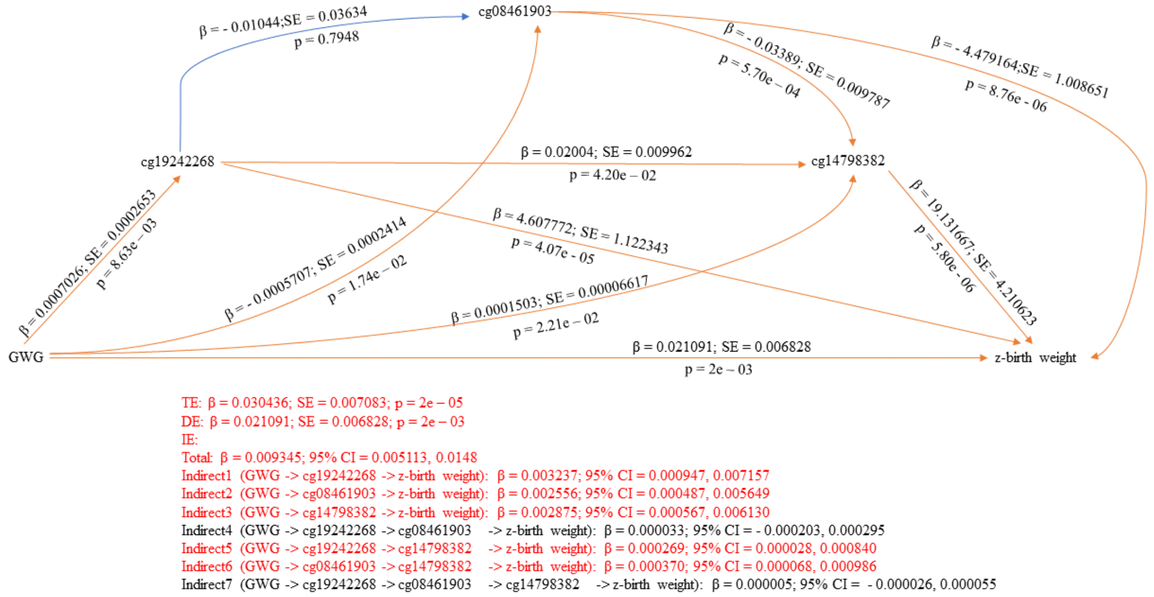


**Figure 2: Serial multiple mediator model as per the OLS bootstrap approach with candidate CpG sites as mediators of the association between GWG and offspring's z-birth weight**

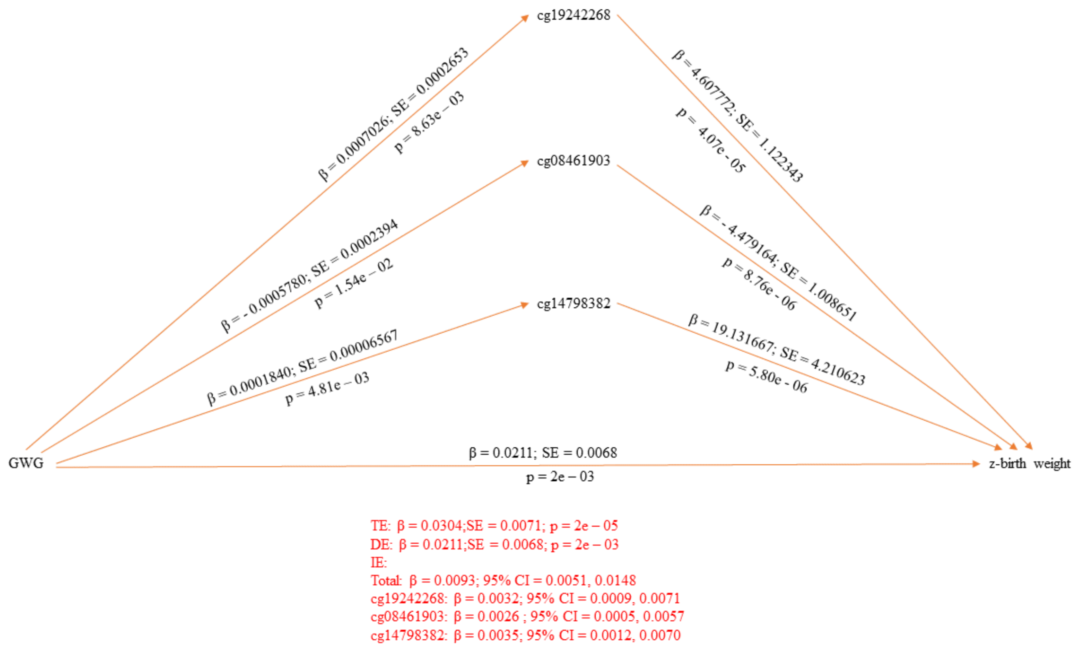


**Figure 3: Parallel multiple mediator model as per the OLS bootstrap approach with candidate CpG sites as mediators of the association between GWG and offspring's z-birth weight**

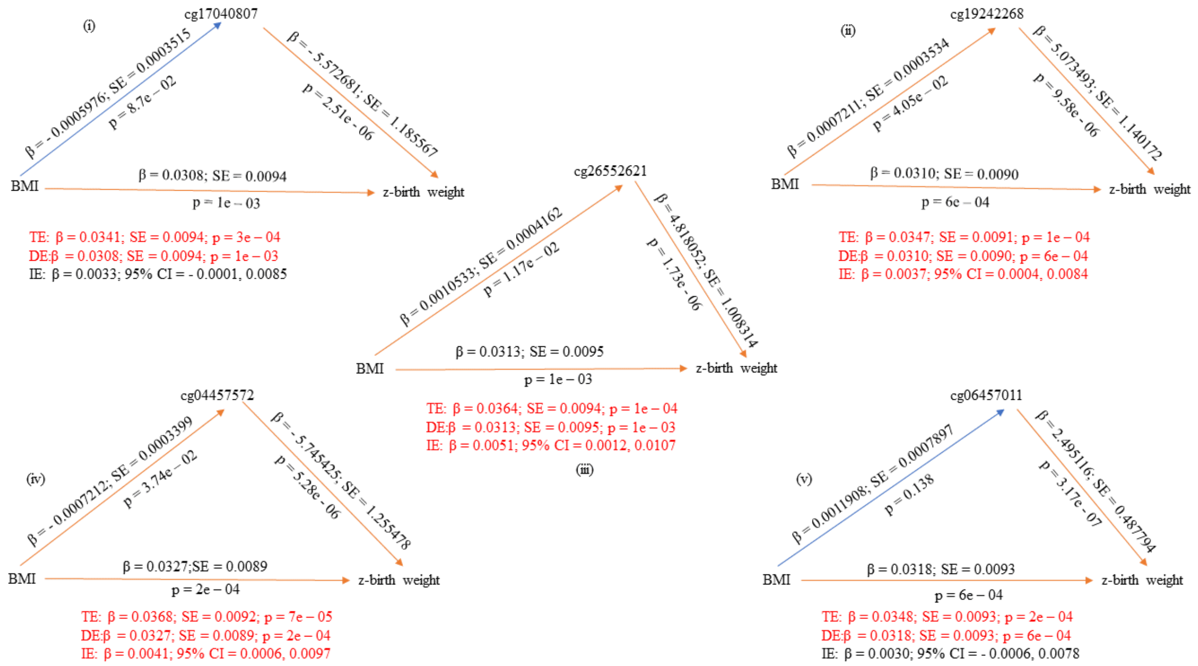


**Figure 4: Single mediator models as per the robust bootstrapped approach with candidate CpG sites as mediators of the association between maternal BMI at the beginning of pregnancy and z-birth weight**

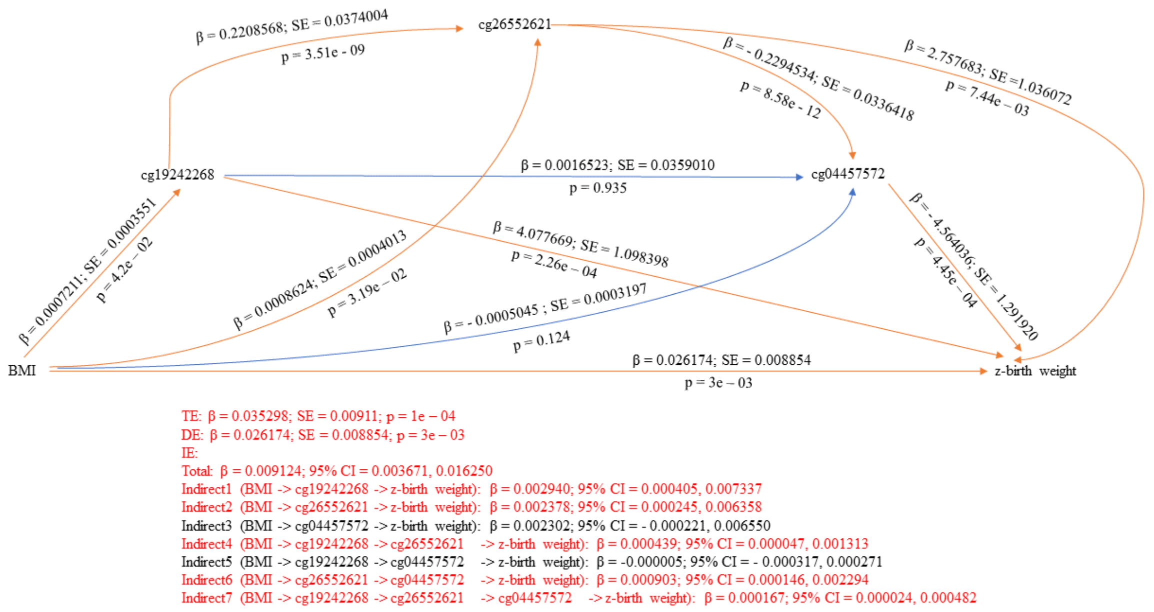


**Figure 5: Serial multiple mediator model as per the robust bootstrapped approach with candidate CpG sites as mediators of the association between maternal BMI at the beginning of pregnancy and z-birth weight**

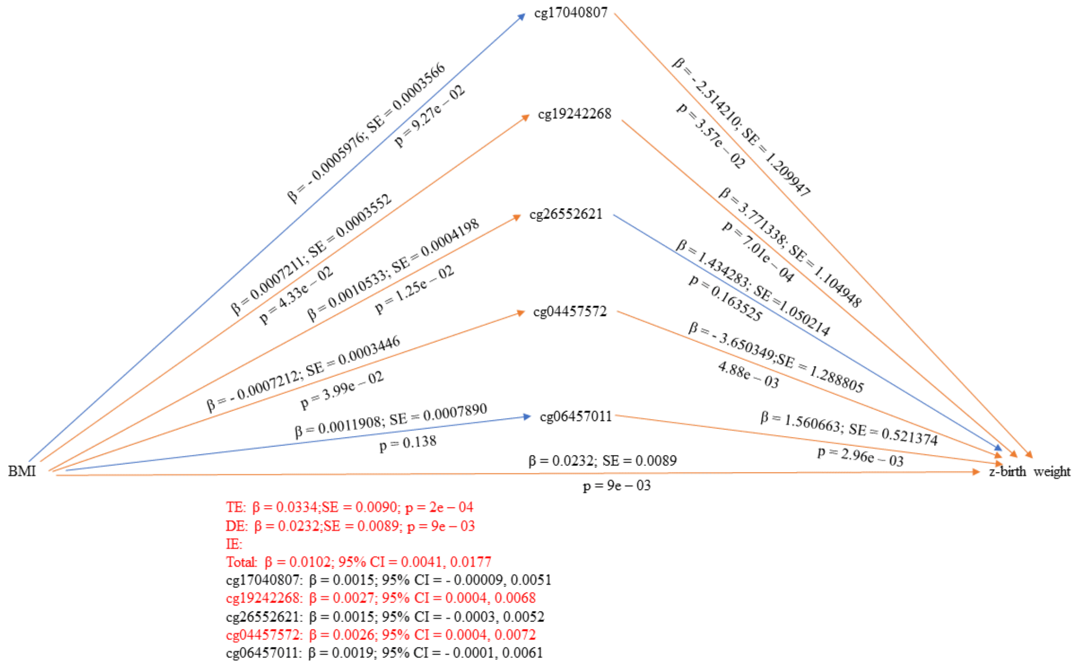


**Figure 6: Parallel multiple mediator model as per the robust bootstrapped approach with candidate CpG sites as mediators of the association between maternal BMI at the beginning of pregnancy and z-birth weight**

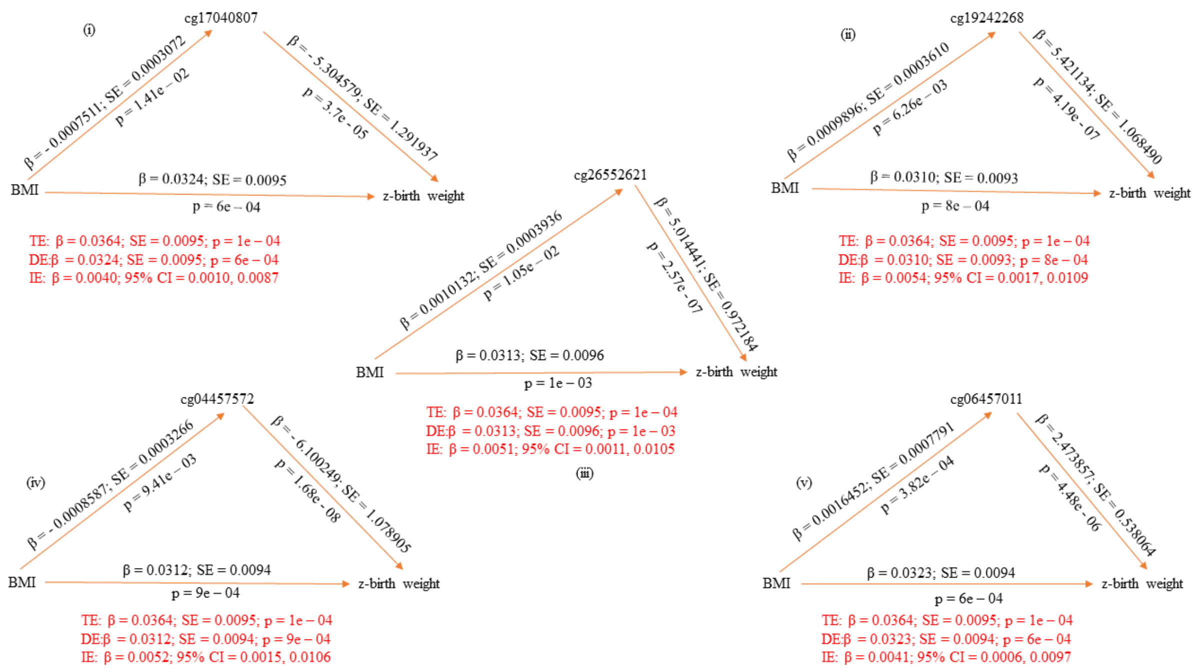


**Figure 7: Single mediator models as per the OLS bootstrap approach with candidate CpG sites as mediators of the association between maternal BMI at the beginning of pregnancy and z-birth weight**

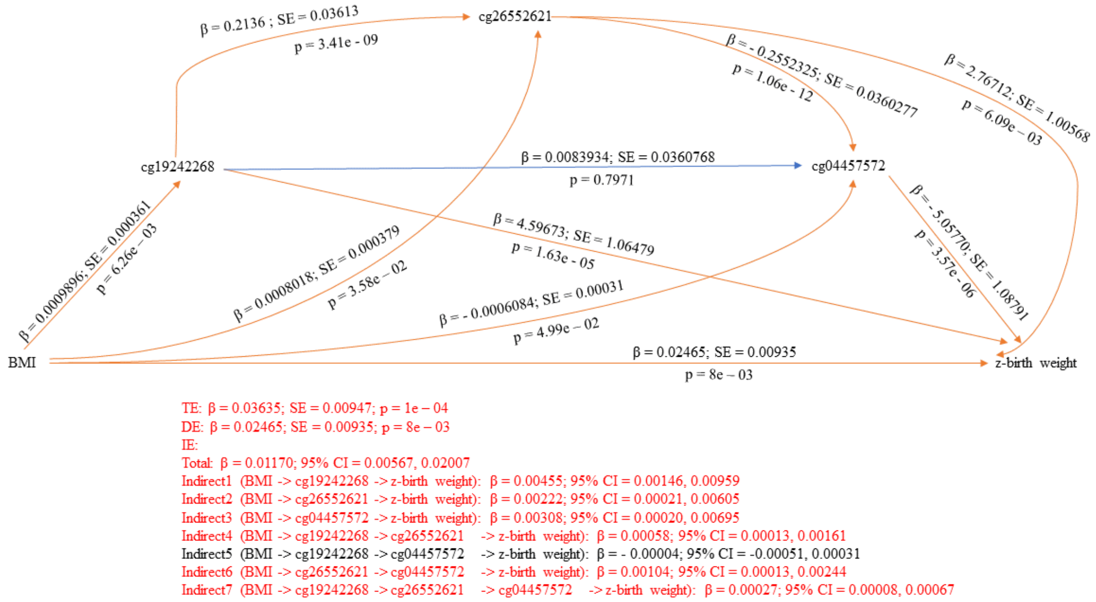


**Figure 8: Serial multiple mediator model as per the OLS bootstrap approach with candidate CpG sites as mediators of the association between maternal BMI at the beginning of pregnancy and z-birth weight**

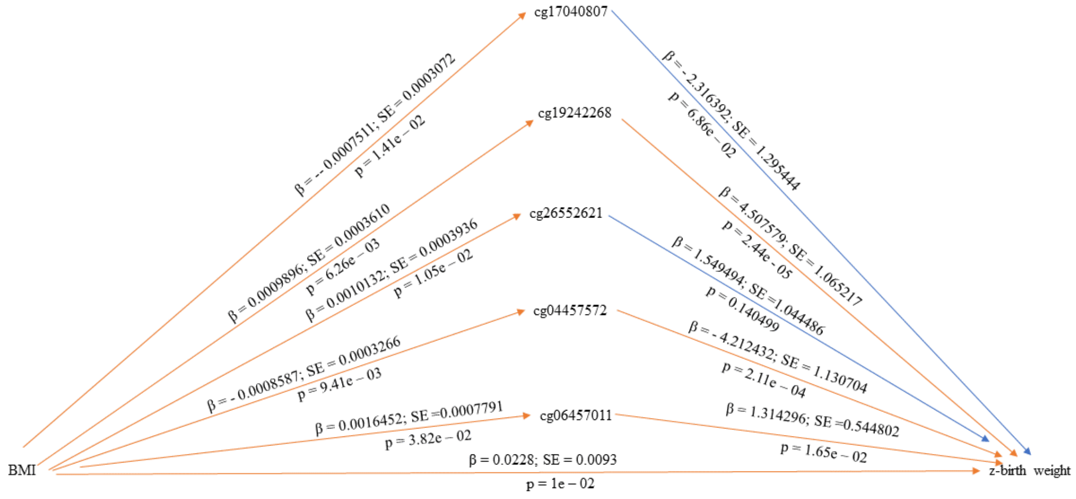


TE: β = 0.0364;SE = 0.0095;p = 1e - 04
DE: β = 0.0228;SE = 0.0093;p = 1e - 02
IE:
Total: β = 0.0136; 95% CI = 0.0069, 0.0224
cg17040807: β = 0.0017; 95% CI = - 0.00001, 0.0050
cg19242268: β = 0.0045; 95% CI = 0.0014, 0.0093
cg26552621: β = 0.0016; 95% CI = - 0.0002, 0.0052
cg04457572: β = 0.0036; 95% CI = 0.0009, 0.0081
cg06457011: β = 0.0022; 95% CI = 0.0002, 0.0064

**Figure 9: Parallel multiple mediator model as per the OLS bootstrap approach with candidate CpG sites as mediators of the association between maternal BMI at the beginning of pregnancy and z-birth weight**

**Supplementary material 9: Findings from previous studies on the association of candidate CpG sites with markers of obesity published on the EWAS Catalog**

| CpG mediator | Previous findings (EWAS Catalog*) |
|---|---|
| cg19242268 | (1) Birth weight EWAS meta-analysis of all ancestries: Küpers et al, 2019; PMID: 31015461<br>(2) Birth weight EWAS meta-analysis of European ancestry only: Küpers et al, 2019; PMID: 31015461<br>(3) Childhood growth trajectories from birth to late adolescence: Mulder et al, 2021; PMID: 33450751 |
| cg21516291 | (1) Birth weight EWAS meta-analysis of all ancestries: Kupers et al, 2019; PMID: 31015461<br>(2) Birth weight EWAS meta-analysis of European ancestry only: Kupers et al, 2019; PMID: 31015461<br>(3) Incident type 2 diabetes: Hillary et al, 2023; PMID: 37410739<br>(4) Childhood growth trajectories from birth to late adolescence: Mulder et al, 2021; PMID: 33450751 |
| cg17040807 | (1) Birth weight EWAS meta-analysis of all ancestries: Kupers et al, 2019; PMID: 31015461 |
| cg26552621 | (1) NOG protein levels (obesogenic): Gadd et al 2022; PMID: 35945220<br>(2) Omega 6 fatty acids: Battram et al; PMID: NA<br>(3) Childhood growth trajectories from birth to late adolescence: Mulder et al, 2021; PMID: 33450751 |
| cg04457572 | (1) Incident type 2 diabetes: Hillary et al, 2023; PMID: 37410739<br>(2) ADIPOQ protein/adiponectin levels (regulate fat metabolism and insulin sensitivity): Gadd et al, 2022; PMID: 35945220<br>(3) Growth trajectories from birth to early childhood: Xu et al, 2017; PMID: 28056824<br>(4) Childhood growth trajectories from birth to late adolescence: Mulder et al, 2021; PMID: 33450751 |
| cg06457011 | (1) Fasting insulin: Liu et al, 2019; PMID: 31197173<br>(2) Estimated degree of fatty acid saturation levels: Battram et al; PMID: NA<br>(3) Childhood growth trajectories from birth to late adolescence: Mulder et al, 2021; PMID: 33450751 |
| cg14798382 | (1) Childhood growth trajectories from birth to late adolescence: Mulder et al, 2021; PMID: 33450751 |
| cg08461903 | (1) Childhood growth trajectories from birth to late adolescence: Mulder et al, 2021; PMID: 33450751 |

* **https://www.ewascatalog.org/**
